# Urethral Morphology and Support Associated with Urinary Symptoms after Vaginal Surgery with and without Midurethral Sling

**DOI:** 10.64898/2026.05.17.26353431

**Authors:** Shaniel T. Bowen, Pamela A. Moalli, Heidi S. Harvie, Charles R. Rardin, Michael E. Hahn, Alison C. Weidner, Holly E. Richter, Tasha Serna-Gallegos, Donna Mazloomdoost, Amaanti Sridhar, Marie G. Gantz, the NICHD Pelvic Floor Disorders Network

## Abstract

**Background:** Midurethral sling placement is often performed during prolapse repair to treat or prevent stress urinary incontinence. However, some women experience persistent or new-onset stress or urgency urinary incontinence after surgery. It is unclear how prolapse repair, with or without a concomitant midurethral sling, alters urethral morphology and support, and how these changes relate to urinary continence outcomes.

**Objectives:** To compare postoperative urethral morphology (dimensions, angles, shape) and support (position, mobility) after transvaginal prolapse repair with vs without a concurrent midurethral sling, and to explore associations between postoperative urethral characteristics and urinary outcomes (stress, urgency symptoms).

**Study Design:** This ancillary analysis used magnetic resonance imaging and urinary outcome data from the Defining Mechanisms of Anterior Vaginal Wall Descent Study conducted across 8 clinical sites within the United States Pelvic Floor Disorders Network. Eighty-two women (median age, 65 years) underwent transvaginal prolapse repair (vaginal mesh hysteropexy or vaginal hysterectomy with uterosacral ligament suspension) with or without a concurrent midurethral sling between April 2013 and February 2015. Postoperative imaging at rest and during strain was performed 30-42 months after surgery (or earlier if they chose reoperation) between June 2014 and May 2018. Prolapse recurrence, defined as descent beyond the vaginal introitus during strain, was recorded. The urethra was segmented from postoperative scans to create 3-dimensional models for measuring urethral diameters, length, surface area, volume, angles, shape (principal component scores from a statistical shape model), position, and mobility (rest-to-strain displacement). Preoperative and 24-48-month postoperative urinary continence outcomes were assessed using validated questionnaires: the Urogenital Distress Inventory, Urinary Impact Questionnaire, and the Incontinence Severity Index. Comparisons of urethral and urinary outcomes by (1) midurethral sling and (2) stress urinary incontinence were made using Wilcoxon rank-sum tests, principal component analysis, and multivariate models as appropriate. Associations between urethral and urinary outcomes were evaluated with Spearman’s rank correlation.

**Results:** Forty-six women (22 hysteropexy, 24 hysterectomy) were in the sling group, and 36 (19 hysteropexy, 17 hysterectomy) were in the no-sling group. Among the 48 women without prolapse recurrence (28 sling, 20 no-sling), those with a sling (vs without) had larger urethral dimensions (all P<.03), a more anterior-superior position of the proximal urethra (indicating better bladder neck support) (P=.04), a straighter urethral shape (P=.006), and reported less bothersome postoperative stress incontinence (P=.02). Overall, 14 women (17%) experienced postoperative stress incontinence. Stress urinary incontinence was linked to a more acute proximal urethral sagittal angle (more aligned with axial plane) (P=.01), and a lower proximal urethra position (P=.04) and mid-urethra position (P=.03). Poorer stress and urgency urinary outcomes were associated with a shorter urethral length (P=.01), a more posterior-inferior urethral position (all P<.05), increased “C” or “S”-shaped urethral concavity (P=.008; P=.006), and smaller rest-to-strain displacement of the proximal (P=.03) and distal (P=.009) urethra.

**Conclusions:** Urethral morphology and support differed with concomitant midurethral sling (vs no sling) and stress urinary incontinence after vaginal surgery. Urethral characteristics were also associated with postoperative urinary symptoms. Urethral configuration may influence urinary outcomes and could be considered during prolapse and stress urinary incontinence repairs.

**CONDENSATION PAGE:** *TWEETABLE STATEMENT:* Vaginal prolapse repair with midurethral sling (vs without) was linked to a larger, straighter urethra, and better urinary outcomes. Poorer urinary outcomes were linked to a shorter, “C”- or “S”-shaped urethra.

*SHORT TITLE:* Urethral Morphology Associated with Midurethral Sling and Postoperative Urinary Outcomes

*AJOG AT A GLANCE:* A. Why was this study conducted?
Some women experience urinary incontinence after prolapse repair with or without a concomitant midurethral sling.
It remains unclear how midurethral sling surgery alters urethral morphology and support, and how these changes relate to postoperative urinary symptoms. B. What are the key findings?
This magnetic resonance imaging study compared postoperative urethral characteristics (morphology, support) and urinary outcomes among women after vaginal prolapse repair with and without concomitant midurethral sling.
Among women without prolapse recurrence, midurethral sling (vs no sling) was associated with larger urethral dimensions, a straighter, better-supported urethra, and superior urinary outcomes.
Poorer urinary outcomes were associated with a shorter, curvier urethra and worse urethral support. C. What does this study add to what is already known?
Postoperative urethral morphology and support differ by midurethral sling and were associated with urinary outcomes after vaginal prolapse repair.
Urethral configuration warrants surgical consideration in prolapse and urinary incontinence repairs.

## INTRODUCTION

Urinary incontinence (UI) often occurs alongside pelvic organ prolapse. Placement of a midurethral sling (MUS) is commonly performed during vaginal prolapse repair to treat and prevent stress UI (SUI).^1^ While MUS can effectively alleviate UI symptoms, some patients still experience persistent and new-onset SUI or urgency UI (UUI) after surgery.^2^ Urethral configuration (morphology, support) is known to play an important role in continence mechanisms.^3–8^ Thus, postoperative urethral configuration may influence urinary outcomes following prolapse and SUI repair.^5^ However, it remains unclear how these procedures alter urethral morphology and support, and how surgery-related urethral changes correlate with postoperative urinary outcomes.

Recent imaging studies have demonstrated the feasibility of assessing the relationship between urethral anatomy, urinary symptoms, and MUS surgery.^5–12^ Despite this, most studies have been limited to non-surgical cohorts, focused on urethral support (position, mobility) rather than urethral morphology, or evaluated urethral and urinary outcomes separately.

The <u>De</u>fining <u>M</u>echanisms of <u>A</u>nterior Vagi<u>n</u>al Wall <u>D</u>escent (DEMAND) study provides an ideal surgical cohort and magnetic resonance imaging (MRI) data to investigate the relationship between concomitant MUS placement, urethral configuration, and urinary function outcomes after vaginal prolapse surgery.^1,13^ This study aimed to: (1) compare postoperative urethral morphology and support following transvaginal prolapse repair with and without concomitant MUS and (2) explore associations between urethral characteristics and postoperative urinary outcomes. Based on prior literature,^5–7,9,10^ we hypothesized that (1) MUS (vs no MUS) would be associated with a straighter urethra, better periurethral support, and improved urinary outcomes, and (2) postoperative SUI (vs no SUI) would be associated with a more curved urethra, poorer urethral support, and worse urinary outcomes.

## MATERIALS AND METHODS

### Study Design

This was a secondary analysis of DEMAND, an eight-center supplementary imaging study of the Study of Uterine Prolapse Procedures-Randomized (SUPeR) trial conducted by the Pelvic Floor Disorders Network.^13,14^ DEMAND identified anatomical factors and mechanisms of prolapse recurrence in a subset of women from SUPeR who received treatment for uterovaginal prolapse through either transvaginal sacrospinous hysteropexy using the Boston Scientific Uphold LITE mesh (hysteropexy) or vaginal hysterectomy with uterosacral ligament fixation (hysterectomy).^13^ MUS placement was determined on a case-by-case basis by the surgeon, not per study protocol.^1^ The study design and primary three-year outcomes have been published.^13,15^ Institutional review board approval and participants’ written informed consent were obtained at each site.

### Study Population

A subset of 88 SUPeR participants enrolled in DEMAND was screened for eligibility for this ancillary study. All inclusion and exclusion criteria are listed in **Supplemental Table 1**. During the primary DEMAND study, postoperative pelvic MRIs were collected 30-42 months after surgery (or earlier if participants chose reoperation within 30 months).

Baseline (preoperative) and 24-48-month (on average) follow-up patient characteristics were obtained from SUPeR, including demographics, medical history, Pelvic Organ Prolapse Quantification (POP-Q) measures, and patient-reported outcomes related to urinary function and condition-specific quality of life.

### Imaging Protocol

Before imaging, participants completed 6-8 training visits on how to strain maximally. During the MRI exam, multiplanar T2-weighted images were acquired with participants in the supine position at rest (with prolapse reduced), maximal strain (from 3-4 attempts to achieve the greatest vaginal descent), and recovery (post-strain rest without prolapse reduced). Imaging was performed using 3T scanners with a pelvic phased array coil. The initial rest scan served as a standard reference position for measuring urethral mobility, while the final rest scan (recovery) provided measurements of the physiological urethral configuration (morphology, position). MRIs were imported into 3D Slicer (v4.10, www.slicer.org) and aligned using a 3D pelvic coordinate system (based on the ischial spines and pubic symphysis), with the anterior-posterior (Y) and superior-inferior (Z) axes defining the midsagittal plane.^16^

### Prolapse Recurrence Definition

In the midsagittal plane of the strain MRI, prolapse recurrence was classified as vaginal wall protrusion extending beyond the line between the external urethral meatus and the anterior margin of the perineal body.

### Image Segmentation and 3D Reconstruction

Using 3D Slicer, the urethra—including the muscularis, submucosa, mucosa, and lumen—was manually segmented from the axial recovery MRIs. Two-dimensional slice-by-slice segmentations were stacked to reconstruct 3D surface models of the urethra. Models were then smoothed in Blender (v2.38, Blender Foundation, Amsterdam, The Netherlands) with a smoothing algorithm that minimized volume loss, shape changes, and model-derived measurement errors.^17^ A pelvic 3D model and MRI of a participant with a concomitant MUS are shown in **Figure 1**.

**Figure 1.**
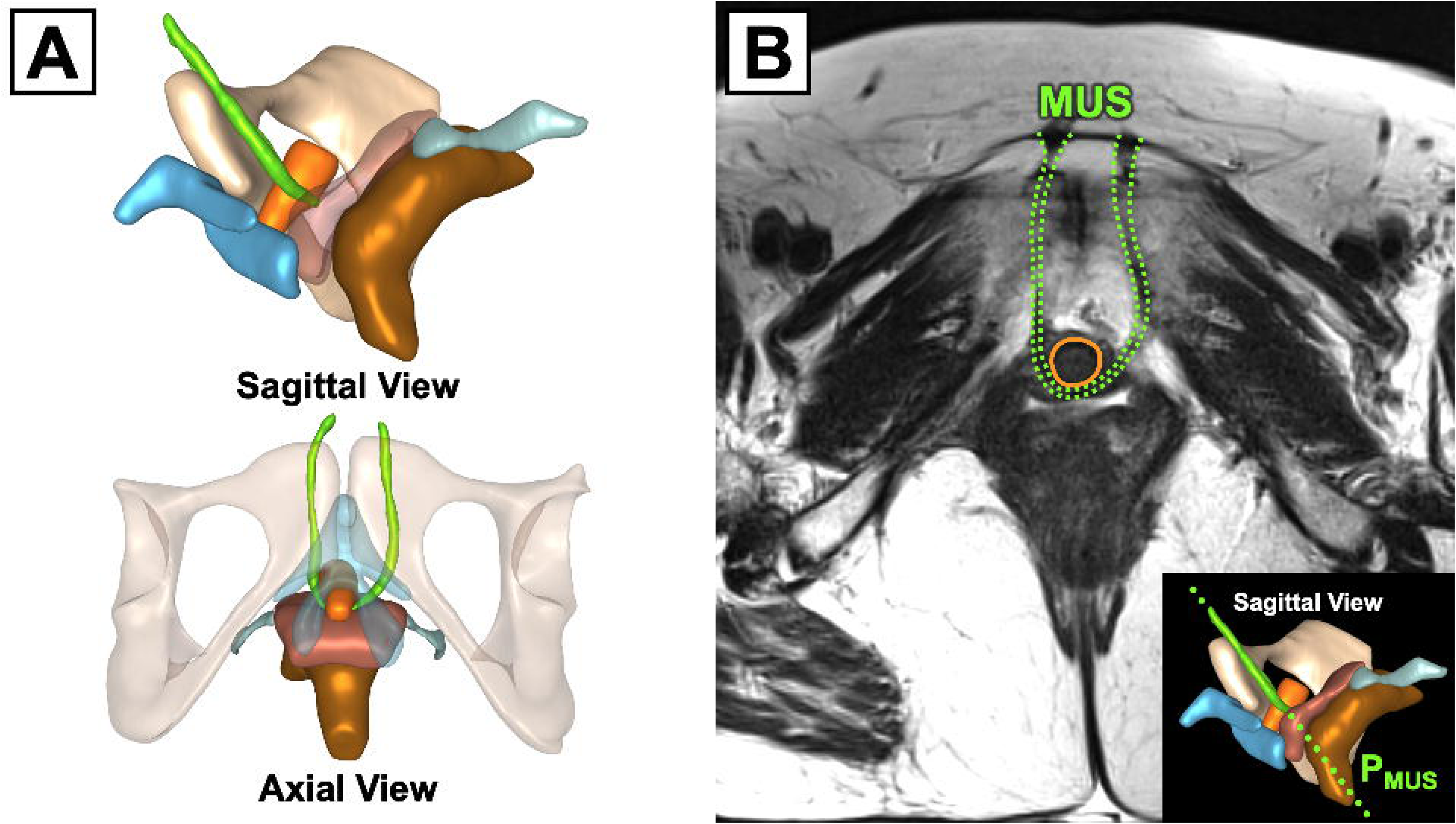
Vaginal Surgery with Concomitant Midurethral Sling. (**A**) Sagittal (top) and axial (bottom) views of a 3D anatomical model, including the pelvis (tan), rectum (brown), uterosacral ligaments (light blue), vagina (red), clitoris (blue), and urethra (orange) of a participant who underwent hysterectomy with concomitant midurethral sling (MUS, green). (**B**) Axial T2-weighted image of the same participant in the plane of the MUS (P_MUS_, green dotted line). Appreciate the “U”-shaped presentation of the retropubic MUS (green dotted line) behind the mid-urethra (orange solid line), with its arms lying approximately 2 cm from the midsagittal plane (pubis symphysis) and inserting into the lower abdominal wall. Note the location of the MUS and its proximity to neighboring pelvic organs (vagina, clitoris) and neurovasculature. Abbreviations: MUS, midurethral sling; P_MUS_, plane of the midurethral sling.

### Urethral Measures

#### Urethral Morphology

Smoothed urethral models were exported to Mathematica (v12.2, Wolfram Research, Champaign, IL) to apply a morphometry algorithm that performed model-based measurements using anatomical landmark detection. Urethral morphology measures were assessed at recovery (rest without prolapse reduced).

##### *Dimensions* (**Figure 2A**)

Urethral models were repeatedly sliced along the axial direction every 1.5 mm from the distal to the proximal urethra. For each urethral slice, the centroid and margins (lateral, anterior, and posterior edges) were identified. The resulting set of centroidal and marginal points defined the urethral centerline and the lateral and anterior-posterior edges, respectively. Then, five equally spaced points along these lines marked the locations of the proximal, mid-proximal, mid-, mid-distal, and distal urethra. At each location, the distance between the anterior-posterior margins measured the anterior-posterior diameter, and the distance between the lateral margins measured the transverse diameter. The length of the centerline determined the urethral length. The surface model was used to compute the urethral surface area and volume.

**Figure 2.**
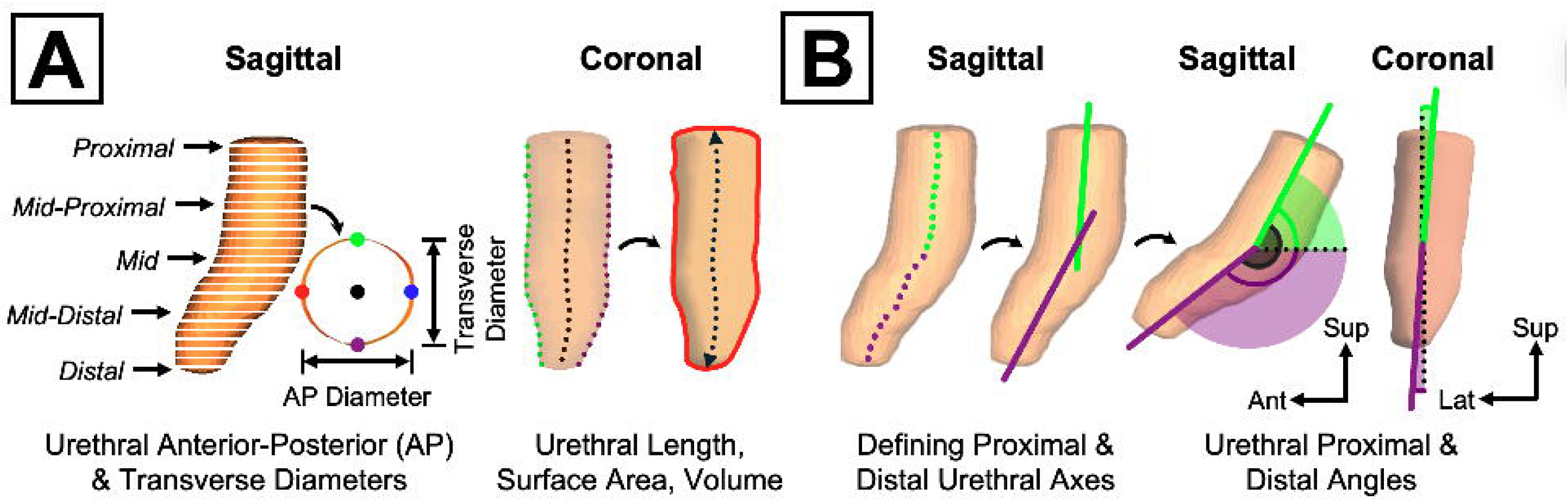
Urethral Dimensional and Angulation Analysis. (**A**) **Urethral Dimensions.** The 3D urethral model (orange) was sliced along the axial direction, where for each slice, the center (black point), anterior (red point), posterior (blue point), and lateral (green and purple points) landmarks were identified. From these landmarks, the anterior-posterior (AP) and transverse diameters were calculated at five equidistant locations along the urethra. The resulting urethral centerline (black dotted line) was used to compute the urethral length (black dotted double arrow), while the 3D surface model was used to measure the urethral cross-sectional area and volume (red solid outline). (**B**) **Urethral Angles.** The upper half (green points) and lower half (purple points) of the urethral centerline defined the proximal urethral axis (green solid line) and distal urethral axis (purple solid outline). The sagittal angles (green arcs) and coronal angles (purple arcs) of—and between (gray arc)—the proximal and distal urethra were calculated relative to the anterior-posterior and superior-inferior axes of the 3D pelvic coordinate system (straight black arrows). Abbreviations: Ant, anterior; AP, anterior-posterior; Lat, lateral; Sup, superior.

##### *Angles* (**Figure 2B**)

The urethral centerline was halved to delineate the proximal and distal urethra. Then, a line of best fit was calculated for each half to define the proximal and distal urethral axes. For each axis, the sagittal and coronal angles relative to the anterior-posterior and superior-inferior axes of the 3D pelvic coordinate system were computed.

##### *Shape* (**Figure 3**)

Urethral shape was analyzed using statistical shape modeling, a method for measuring geometric variability within a population.^18^ First, urethral models were aligned with an iterative closest point algorithm. Next, corresponding points were established by warping a template urethral model (based on the population average) into each patient-specific model using Deformetrica (v4.3).^19^ This process created new patient-specific urethral models composed of points sharing the same anatomical landmarks. These models were then further aligned and scaled in Mathematica. A principal component (PC) analysis was performed on the coordinates of the corresponding points to identify PCs, each representing a portion of the total shape variation within the study group. Significant PCs (i.e., PCs explaining more shape variance than noise, given the dataset size) were identified using a Monte Carlo-based parallel analysis. Each PC includes a set of PC scores that indicate participants’ positions along the shape variation described by the PC, visualized with a heat map. The heat map shows the magnitude (absolute distance between corresponding points) and location of shape differences among urethral models three standard deviations (SDs) from the population mean, with redder colors indicating larger shape differences.

**Figure 3.**
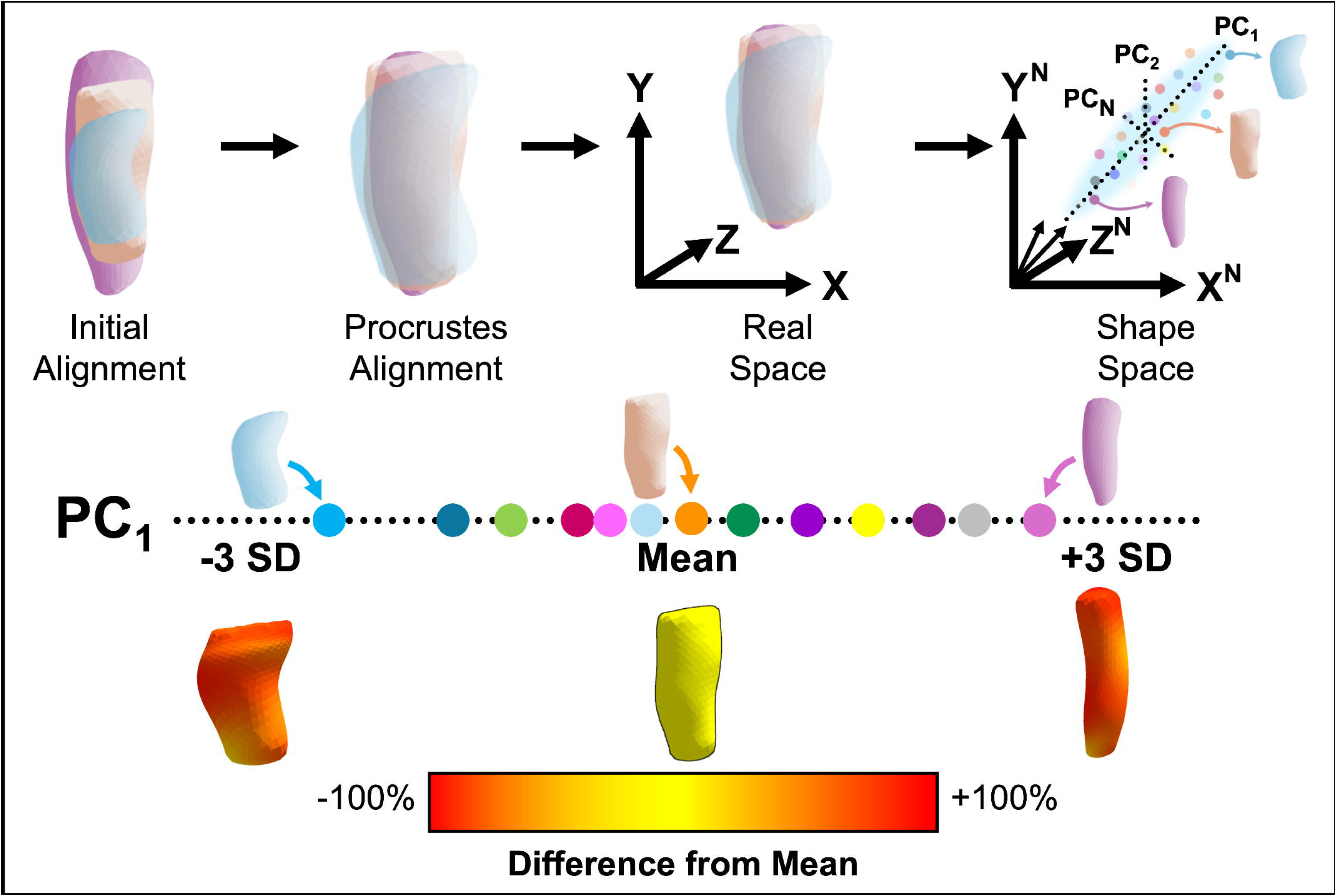
Urethral Statistical Shape Analysis. Visualization of the statistical shape analysis workflow in which 3D urethral models were aligned, scaled, and converted from real space to shape space prior to performing a principal component (PC) analysis on the point coordinates. Each significant PC comprised a set of PC scores that show where individuals lie along the shape variation explained by the PC. To more readily interpret shape differences, a color map was projected onto urethral shapes 3 standard deviations (SDs) from the population mean, with redder colors indicating greater shape differences (normalized, absolute point-to-point distances) from the population mean. Abbreviation: PC, principal component; SD, standard deviation.

#### Urethral Support

Urethral support measures (position, mobility) were computed relative to the pubic symphysis in the midsagittal plane.

##### *Position* (**Figure 4A**)

The location of the proximal, mid-proximal, mid-, mid-distal, and distal urethra was calculated at recovery. The alpha angle—the angle between the anterior and inferior pubic symphysis points and the proximal urethra point—was also measured.

**Figure 4.**
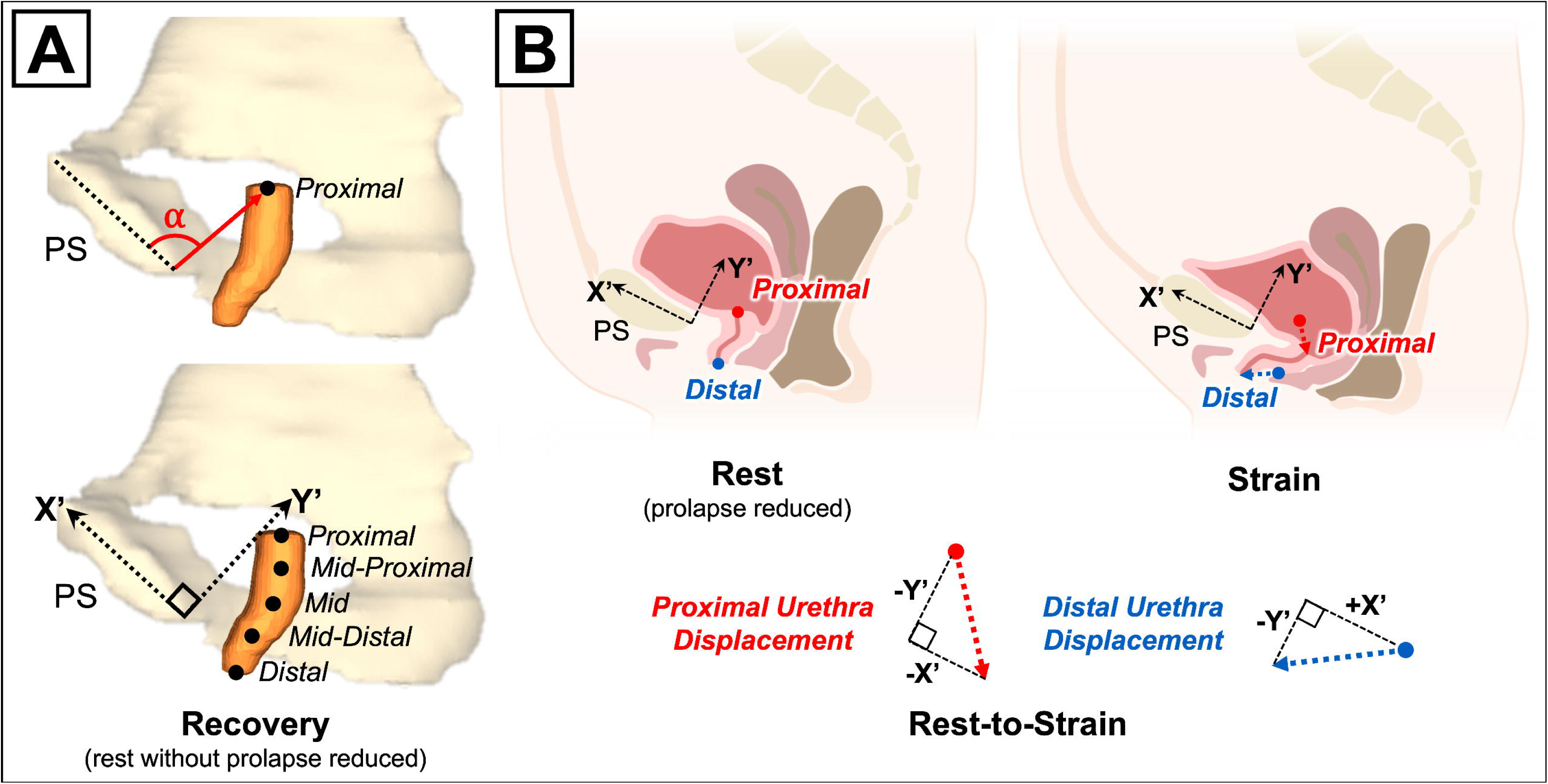
Urethral Position and Mobility Analysis. (**A**) **Urethral Position.** Position of the proximal urethra and four other equidistant urethral locations (black points) at recovery (rest without prolapse reduced) relative to a 2D coordinate system based on the inferior pubic point and midline of the pubic symphysis (PS) (dotted black arrows, X’, Y’). The alpha (α) angle (red arc) also quantified proximal urethral position, defined as the angle between the PS midline (dotted black line) and the proximal urethral point. (**B**) **Urethral Mobility.** The rest (with prolapse reduced, left) to maximal strain (right) displacement (colored arrows) of the bladder neck (red point) and external urethral meatus (blue point) approximated the mobility of the proximal urethra (red dotted arrow) and distal urethra (blue dotted arrow). Abbreviations: PS, pubic symphysis.

##### *Mobility* (**Figure 4B**)

The mobility of the proximal and distal urethra was defined as the magnitude of the rest (with prolapse reduced) to strain displacements of the bladder neck and external urethral meatus.

### Urinary Outcome Measures

Patient-reported outcomes related to urinary function and condition-specific quality of life were evaluated at baseline (preoperative) and follow-up (SUPeR visits at 6, 12, 18, 24, 36, and 48 months postoperatively) using the Urogenital Distress Inventory (UDI-6) Short Form, Urinary Impact Questionnaire (UIQ), and Incontinence Severity Index (ISI). For participants whose MRI exam occurred around 30- and 42-months after surgery, data were imputed from the nearest SUPeR visit to the MRI exam date (e.g., 24-, 36-, or 48-month follow-up visit).

Postoperative SUI status was based on UDI-6 item 3, which asked whether participants experienced urine leakage during coughing, sneezing, or laughing. UDI-6, UIQ, and ISI summary and item scores concerning the degree of UI symptom distress, impact of UI on daily living, and UI severity, respectively, were collected. Higher (worse) scores indicated greater urinary distress, impact, and severity. Additional urinary outcomes included UUI (based on UDI-6 item 2, which asked whether participants experienced urine leakage related to urgency), new or worsening UI, postoperative UI treatment (medication, pelvic floor muscle therapy, percutaneous tibial nerve stimulation), any or bothersome urinary leakage, and post-MUS surgery complications (mesh erosion, mesh exposure, sling revision).

### Sample Size Calculation

The minimum sample size calculated from the primary DEMAND study to detect a moderate effect size (0.5 SD) with 80% power was 40 per vaginal surgery group.^13^

### Statistical Analysis

Baseline and follow-up patient characteristics, postoperative urethral measures (morphology, support), and patient-reported urinary outcomes were summarized as counts (percentages) or medians (interquartile ranges). These were compared by (1) MUS and (2) postoperative SUI status using Fisher’s exact tests and Wilcoxon Rank-Sum tests when appropriate. To isolate the effect of MUS, urethral and urinary outcomes were compared by MUS status among women without prolapse recurrence. The relationship between urethral characteristics and urinary outcomes was assessed using Spearman’s rank correlation. Linear models, incorporating vaginal surgery type, MUS, prolapse recurrence status, their pairwise interactions, and age, were used to evaluate differences in urethral shape (PC scores) between MUS groups.

## RESULTS

### Study Population

Eighty-two of the 88 women from the primary DEMAND study were analyzed: 46 in the MUS group (22 hysteropexy, 24 hysterectomy) and 36 in the No-MUS group (19 hysteropexy, 17 hysterectomy) (**Supplemental Figure 1**). The median (range) age was 65 (47-79) years, with most women being White (82%) and postmenopausal (98%). Forty-eight women (59%) did not experience prolapse recurrence (28 MUS, 20 No-MUS). Based on postoperative SUI status, 14 (17%) were in the SUI group (2 MUS, 12 No-MUS), and 68 (83%) were in the No-SUI group (44 MUS, 24 No-MUS).

Baseline and follow-up participant characteristics by MUS group—overall and among women without prolapse recurrence—are shown in **Supplemental Table 2** and **Table 1**, respectively. Within the no prolapse recurrence group, women with MUS (vs without) had a lower body weight (P=.02), lower body mass index (P=.002), and lower obesity rate (P=.01). Comparisons of patient characteristics by postoperative SUI status are shown in **Table 2**. Women with SUI (vs without) after surgery had a higher rate of cardiovascular disease (P<.001), a lower proportion of MUS repairs (P<.001), and less improvement in anterior vaginal wall support (smaller change from baseline POP-Q Ba) (P=.04).

**Table 1.**
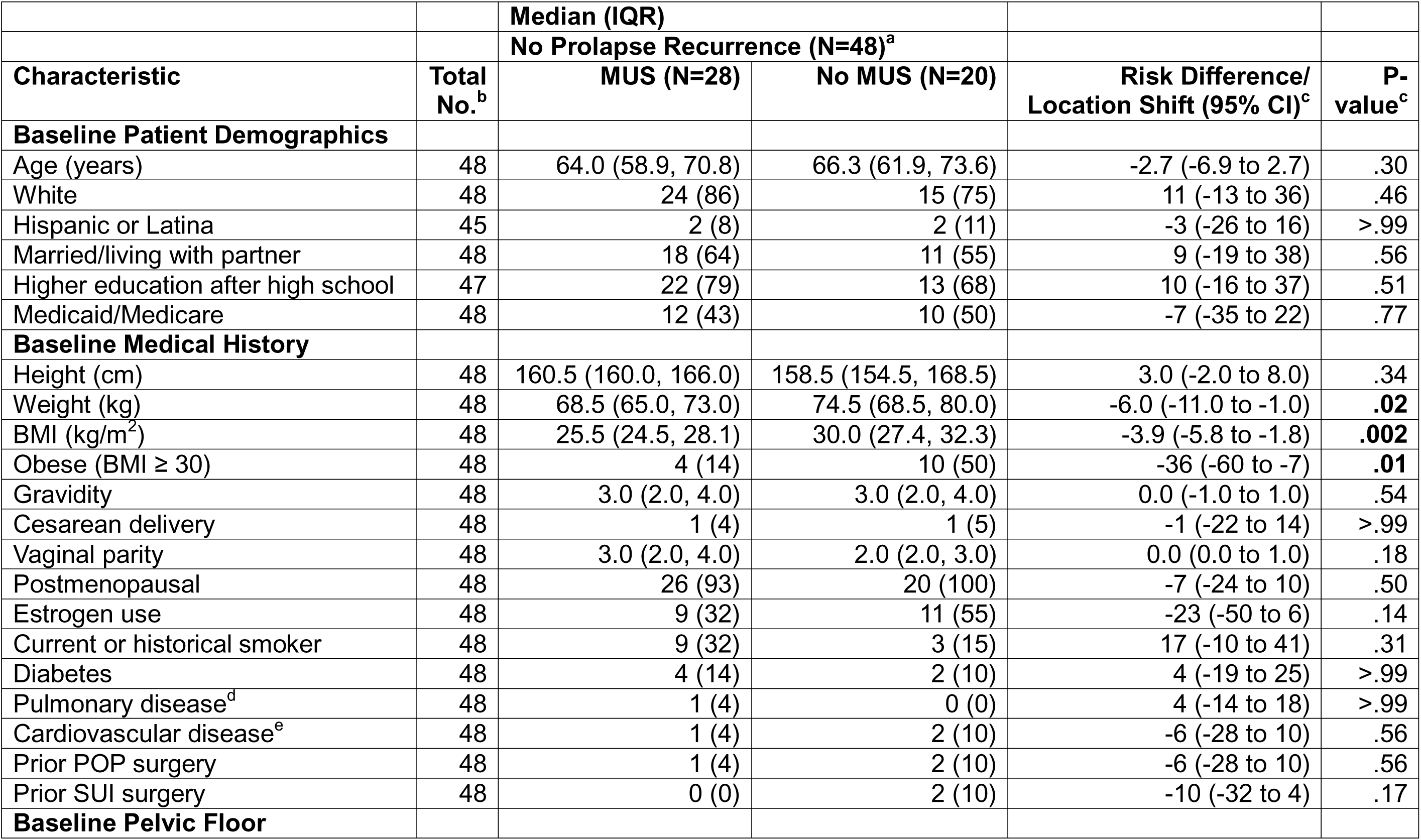
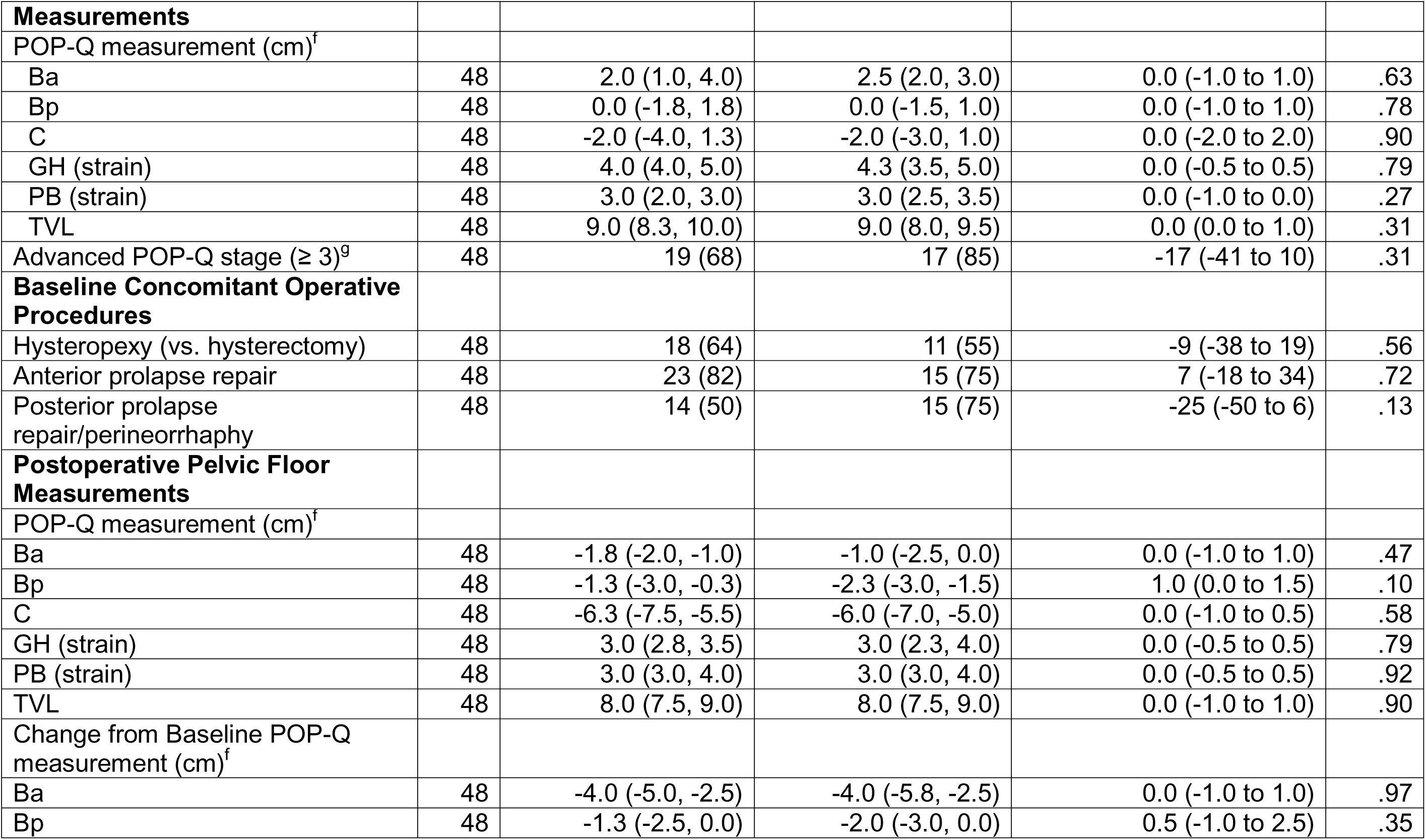
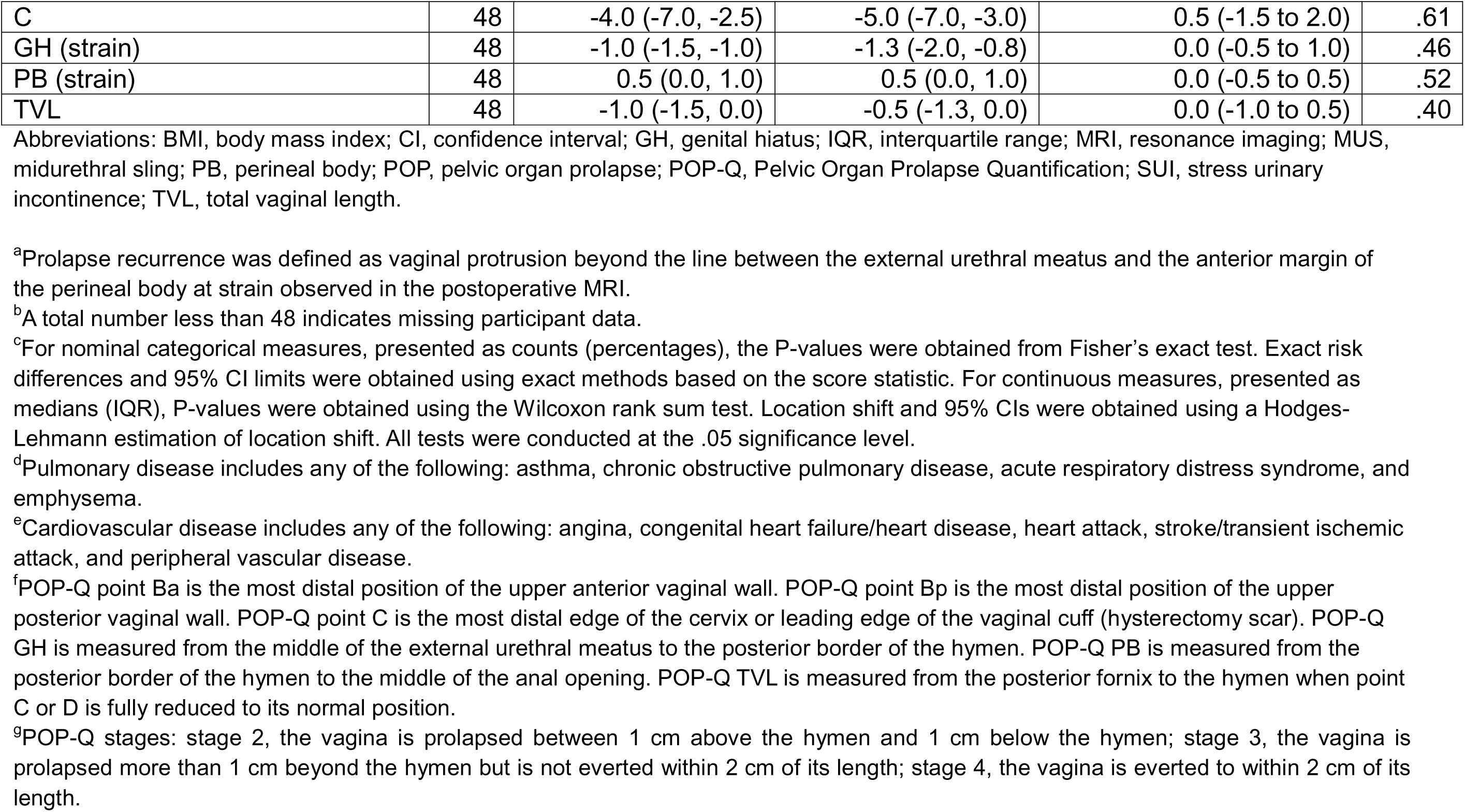
Baseline and Postoperative Demographic and Clinical Characteristics by Midurethral Sling Status among Women without Prolapse Recurrence.

**Table 2.**
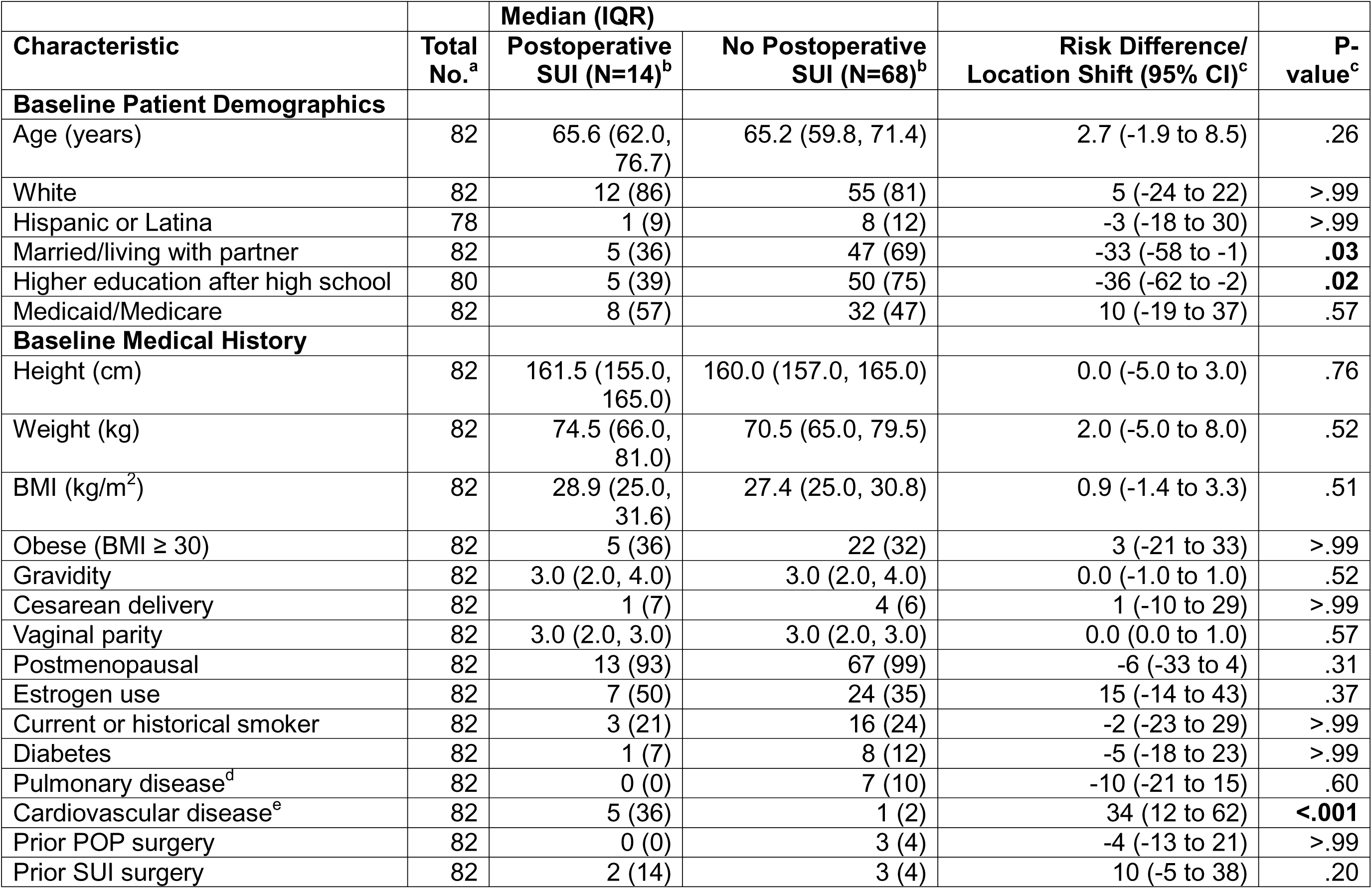
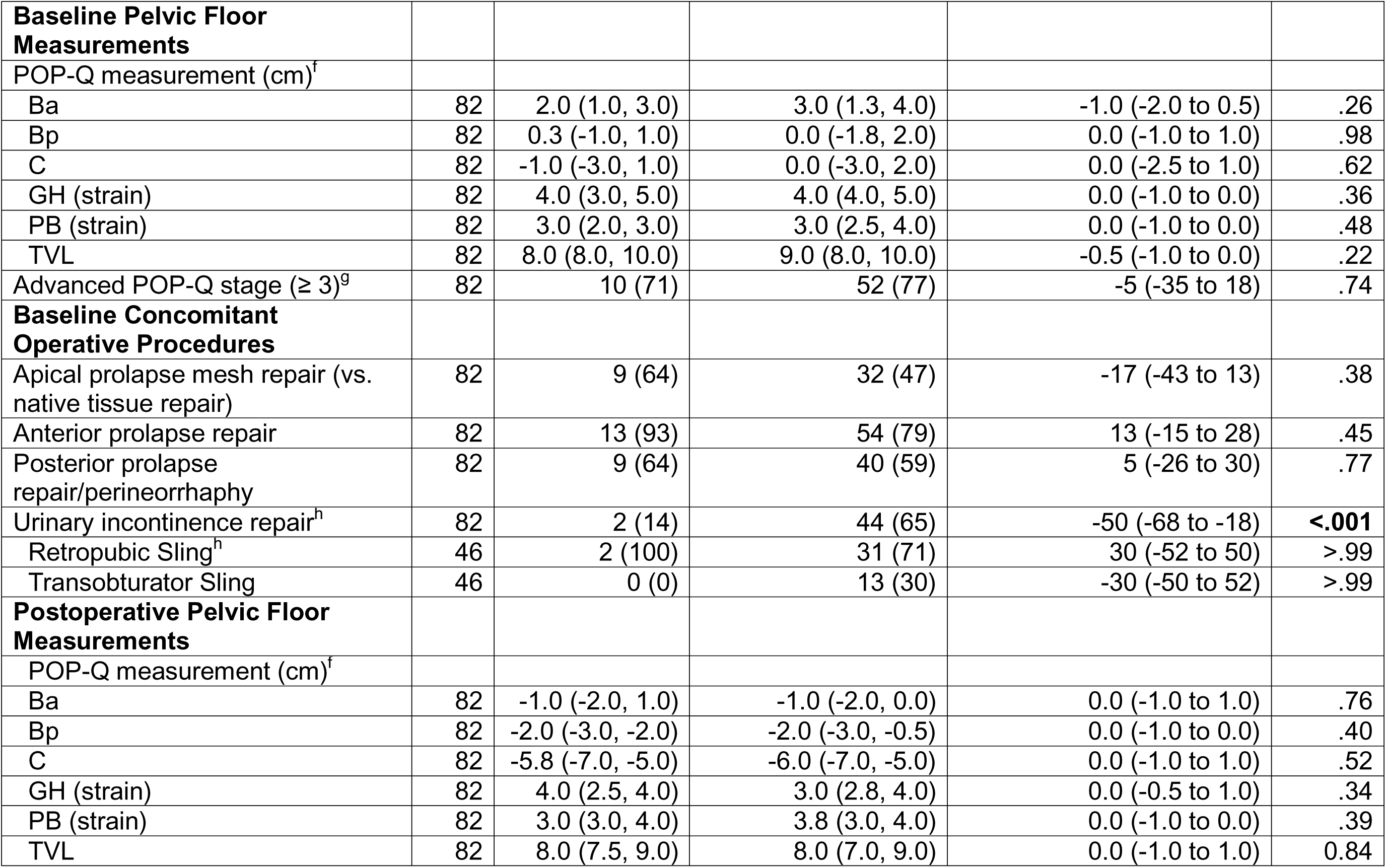
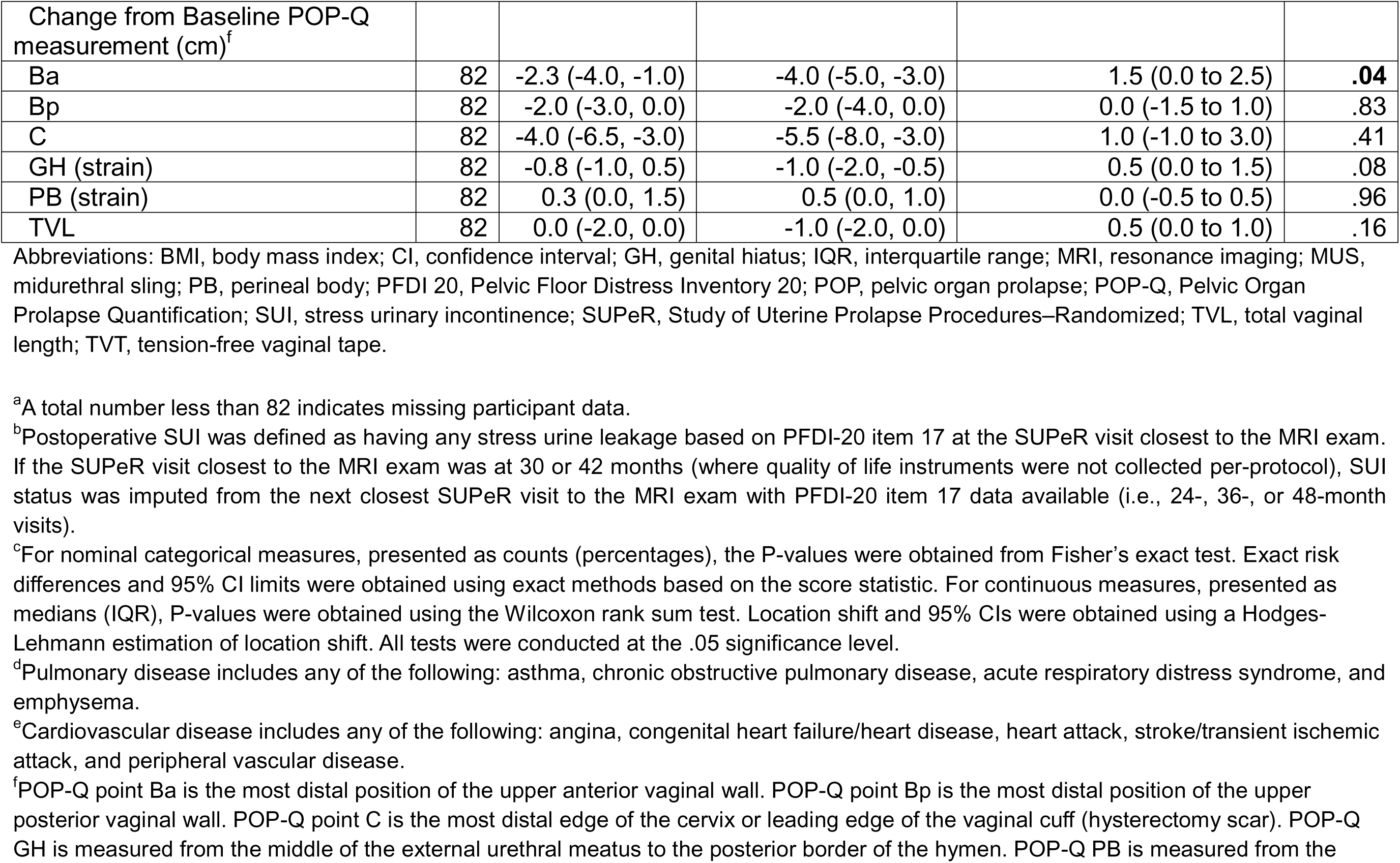
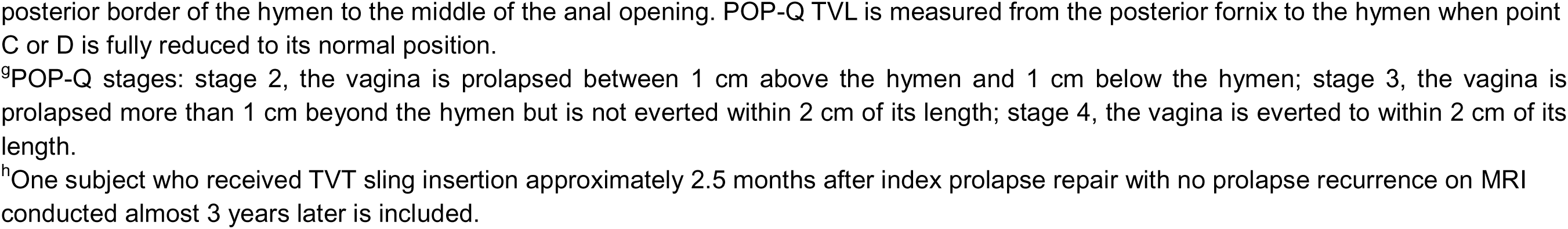
Baseline and Postoperative Demographic and Clinical Characteristics by Postoperative Stress Urinary Incontinence Status.

### Urethral Outcomes

Postoperative urethral characteristics stratified by MUS status across the study population and within the no prolapse recurrence group are shown in **Supplemental Table 3** and **Table 3**, respectively. Among women without prolapse recurrence, the MUS group (vs the No-MUS group) had a larger transverse diameter at the proximal (difference, 1.5 mm; P=.01), mid (difference, 1.1 mm; P=.02), and mid-distal urethra (difference, 1.5 mm; P=.01); a larger surface area (difference, 2.0 cm^2^; P=.01) and volume (difference, 0.8 cm^3^; P=.02); and better proximal urethral support—indicated by a more anterior-superior position of the proximal urethra (difference, 3.0 mm; P=.04) and a smaller alpha angle (difference of -9.7°; P=.04). The statistical shape analysis revealed that among women without prolapse recurrence, women with a MUS exhibited a straighter urethra, while women without a MUS had a more “S”-shaped urethra (PCs 5-6) (P=.006; P=.03) (**Figure 5A/5C**).

**Figure 5.**
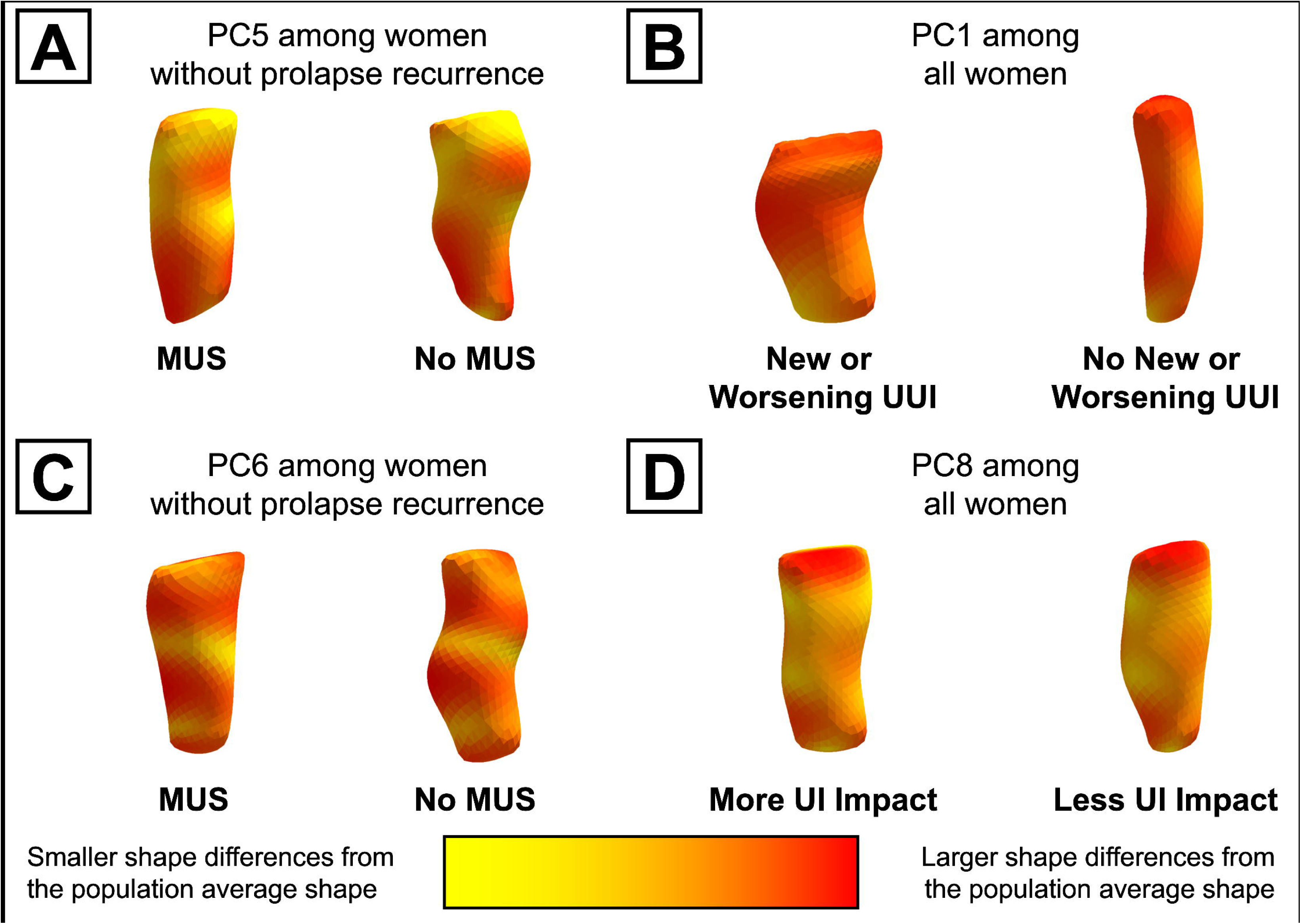
Postoperative Urethral Shape Differences by Midurethral Sling Status and Urinary Outcomes. Urethral shapes three standard deviations from the population average shape for principal components (PCs) 5 (**A**), 1 (**B**), 6 (**C**), and 8 (**D**) that were associated with midurethral sling (MUS) and postoperative urinary incontinence (UI) outcomes, including urge UI (UUI) and impact of UI on daily living (given by the Urinary Impact Questionnaire score). Redder colors indicate larger shape differences from the population mean urethral shape. Shape differences by MUS among women without prolapse recurrence are shown on the left (**A**, **C**), while shape comparisons by urinary outcomes across the entire study population are shown on the right (**B**, **D**). Abbreviations: MUS, midurethral sling; PC, principal component; UI, urinary incontinence; UUI, urge urinary incontinence.

**Table 3.**
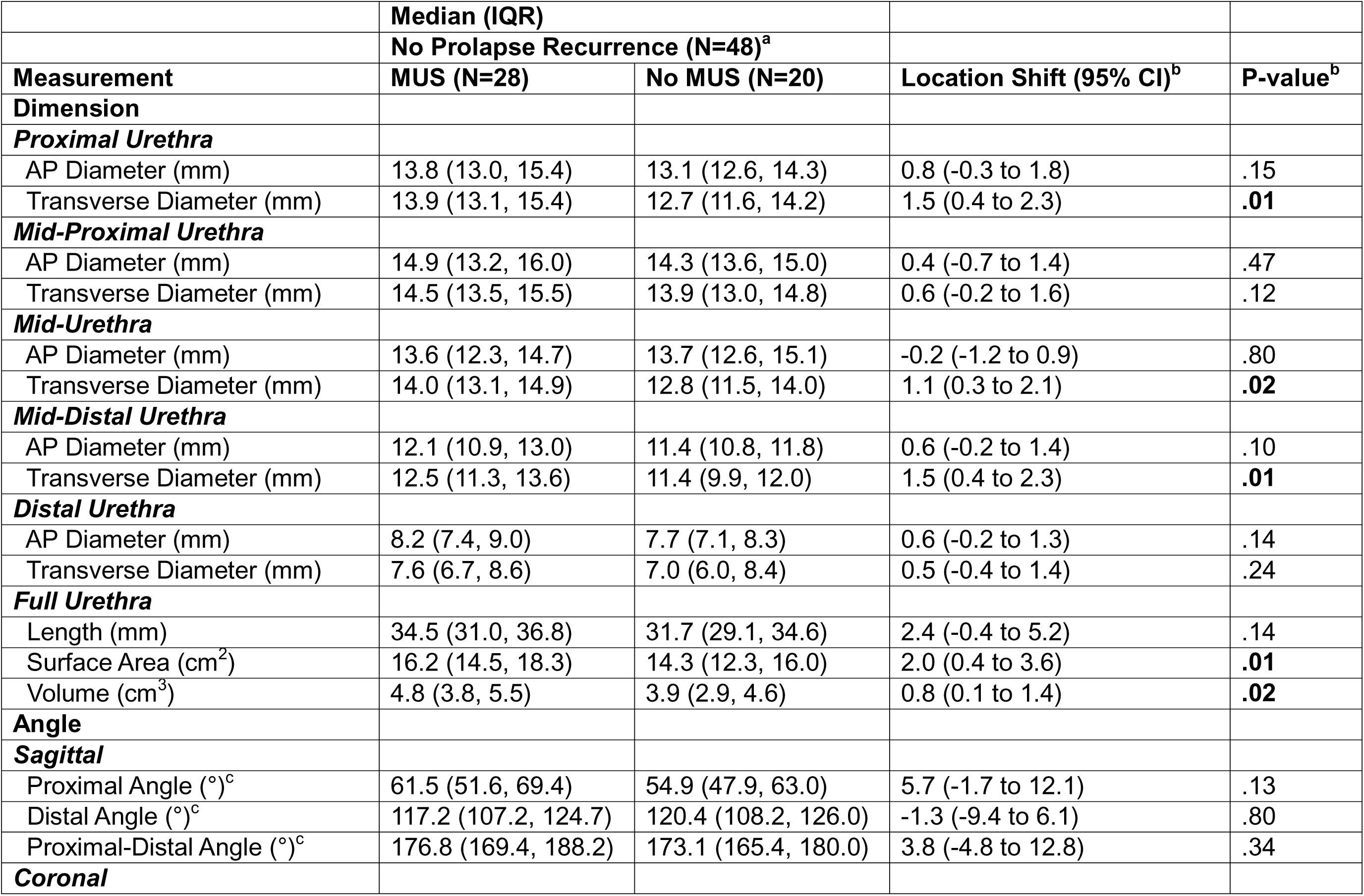
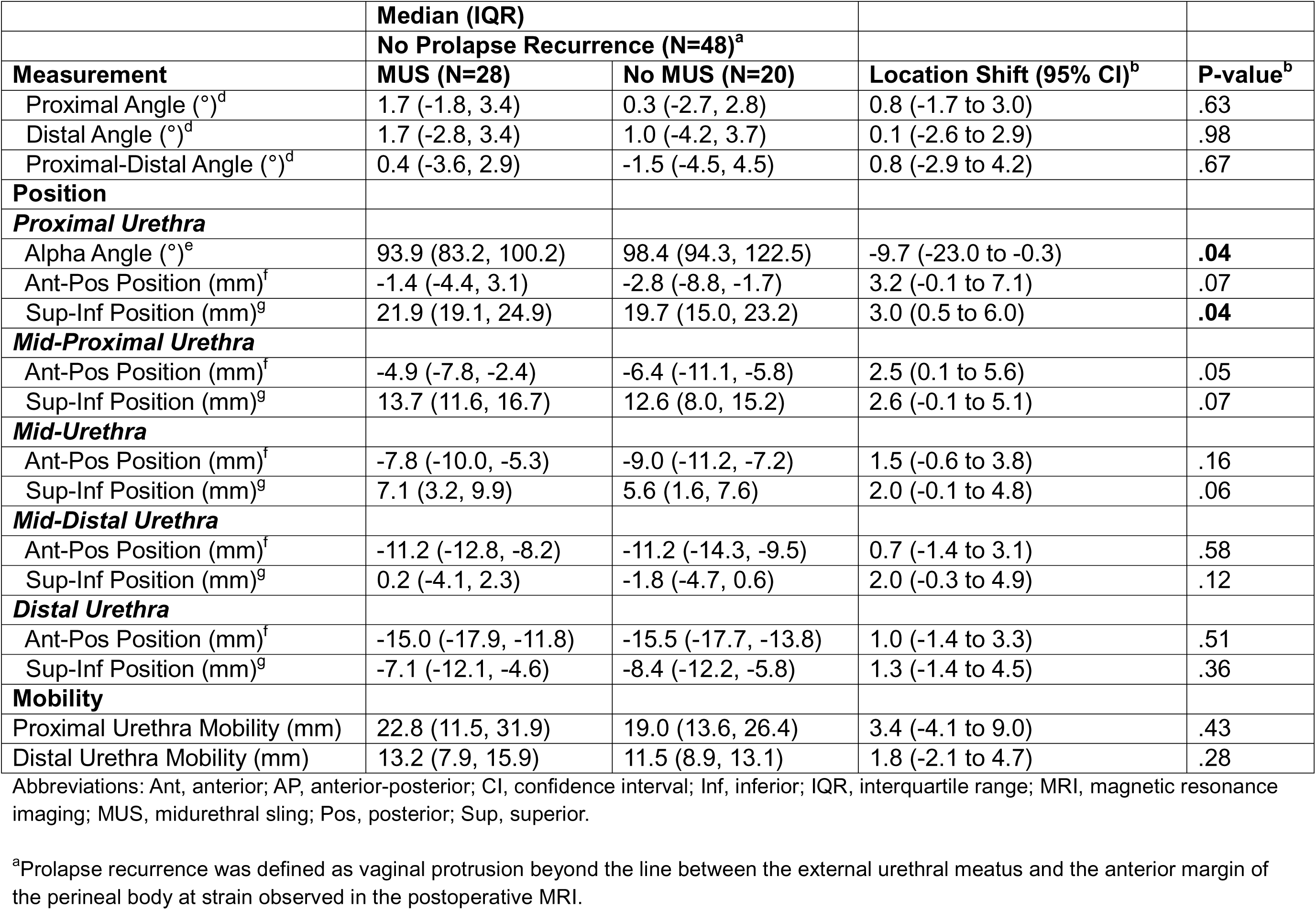
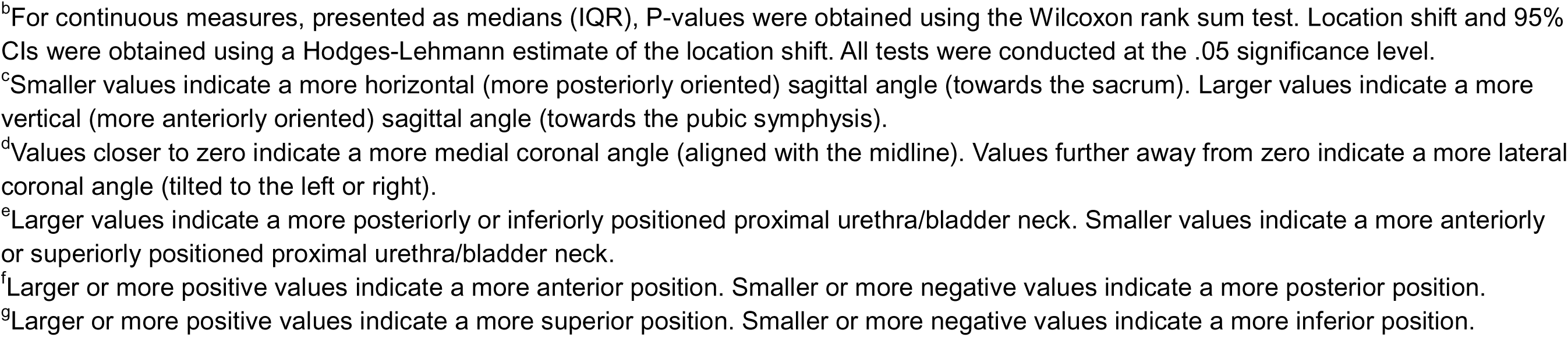
Postoperative Urethral Measurements by Midurethral Sling Status among Women without Prolapse Recurrence.

Group differences in urethral measures based on postoperative SUI status are shown in **Table 4**. Women with SUI (vs without) after surgery had a smaller (more axially oriented) proximal sagittal angle (difference, -8.4°; P=.01) and worse urethral support, indicated by a more posterior-inferior position of the proximal (difference, -3.6 mm; P=.04), mid (difference, -2.8 mm; P=.03), and mid-distal urethra (difference, -2.7 mm; P=.03), along with a larger alpha angle (difference, 12.0°; P=.02). Urethral mobility did not differ by MUS nor postoperative SUI status.

**Table 4.**
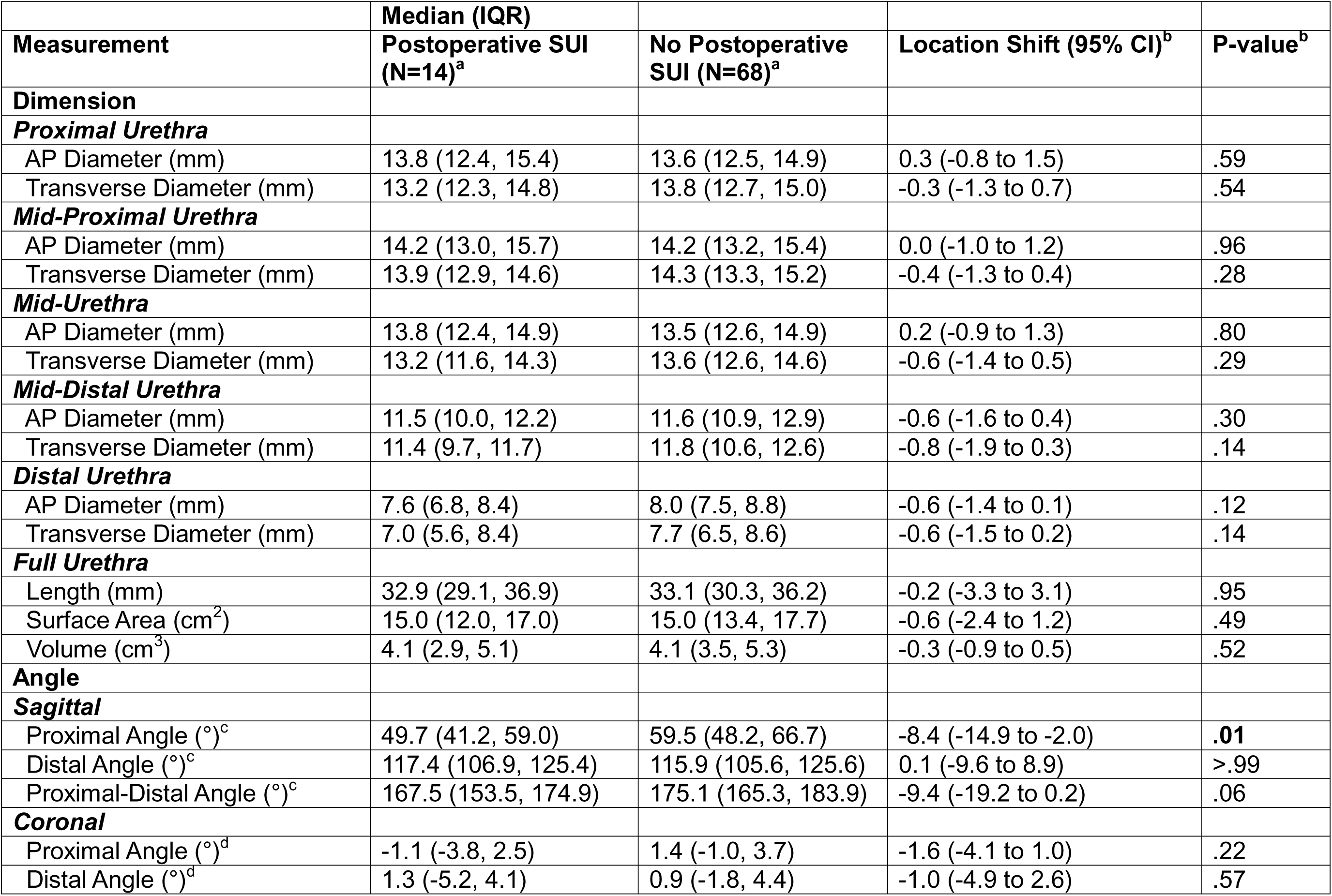
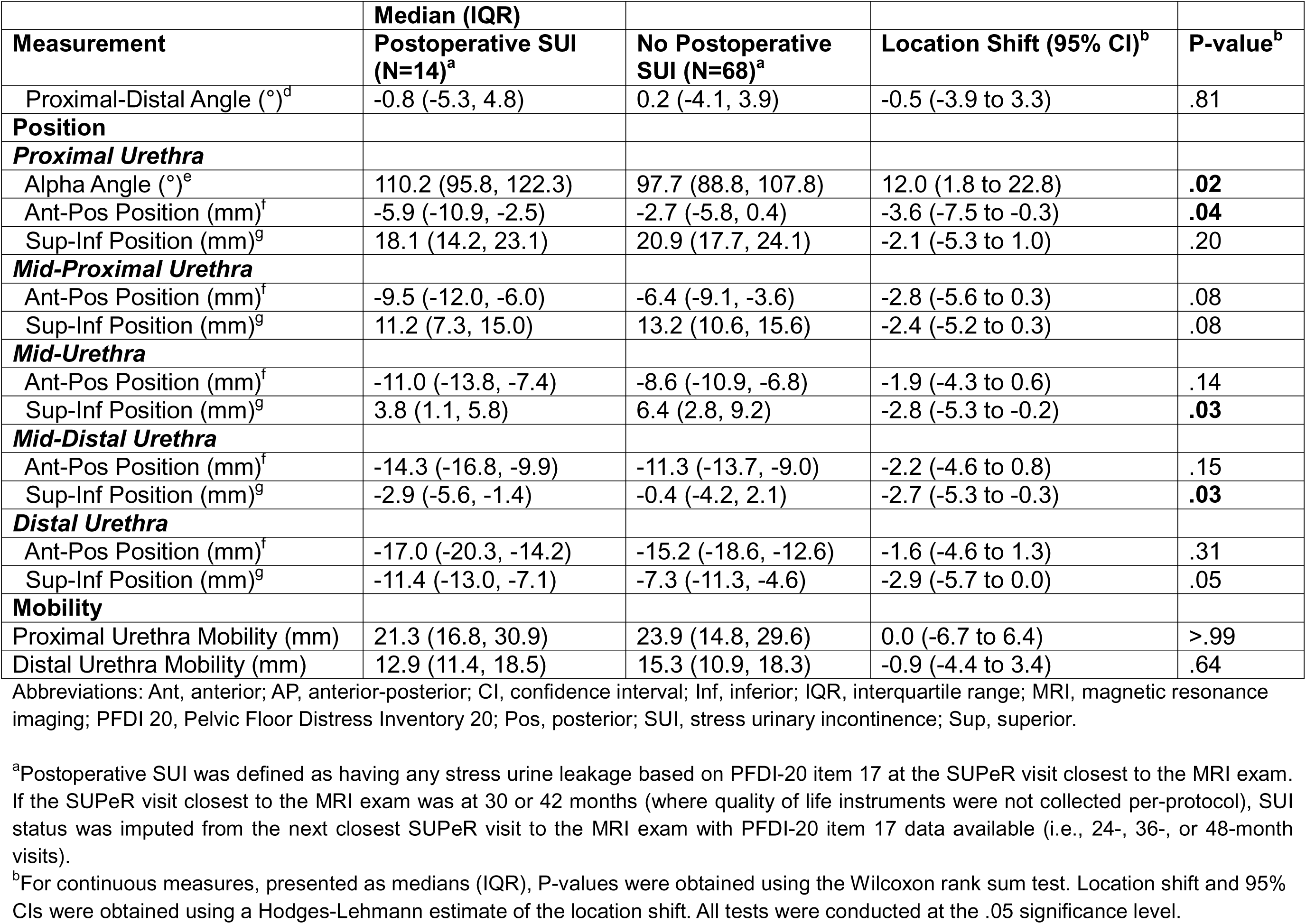
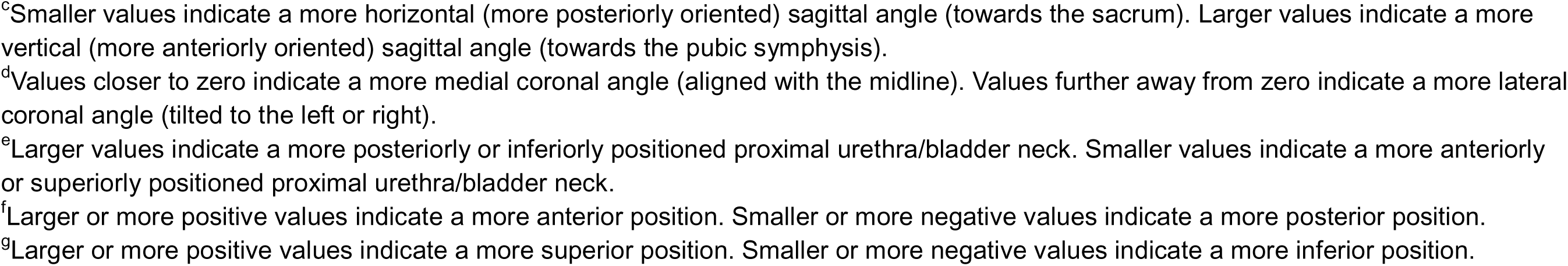
Postoperative Urethral Measurements by Postoperative Stress Urinary Incontinence Status.

### Urinary Outcomes

Comparisons of urinary function outcomes before and after surgery between MUS groups across the study populations and among women without prolapse recurrence are demonstrated in **Table 5**. At baseline, the MUS group (vs No-MUS) had a higher rate of preoperative stress urine leakage (difference, 39%; P<.001) and bother (difference, 37%; P=.002), as well as a higher (worse) ISI score (difference, 2.0; P=.04).

**Table 5.** Baseline and Postoperative Urinary Outcomes by Midurethral Sling Status.

| Urinary Outcome | Total No. | Median (IQR) |  | Risk Difference/<br>Location Shift (95% CI)<sup>a</sup> | P-value<sup>a</sup> |
| --- | --- | --- | --- | --- | --- |
|  |  | MUS (N=46) | No MUS (N=36) |  |  |
| Baseline Urinary Outcomes |  |  |  |  |  |
| Any stress urine leakage<sup>b</sup> | 82 | 32 (70) | 11 (31) | 39 (14 to 58) | <.001 |
| Bothersome stress urine leakage<sup>b</sup> | 82 | 31 (67) | 11 (31) | 37 (12 to 56) | .002 |
| Any urge urine leakage<sup>c</sup> | 82 | 31 (67) | 19 (53) | 15 (-7 to 36) | .25 |
| Bothersome urge urine leakage<sup>c</sup> | 82 | 30 (65) | 19 (53) | 12 (-9 to 34) | .27 |
| Urogenital Distress Inventory (UDI) score | 82 | 47.9 (25.0, 66.7) | 33.3 (18.8, 52.1) | 8.3 (-4.2 to 20.8) | .14 |
| Urinary Impact Questionnaire (UIQ) score | 82 | 9.5 (0.0, 38.1) | 16.7 (0.0, 45.2) | 0.0 (-9.5 to 4.8) | .81 |
| Incontinence Severity Index (ISI) score | 82 | 4.0 (2.0, 8.0) | 2.0 (0.0, 6.0) | 2.0 (0.0 to 3.0) | .04 |
| Postoperative Urinary Outcomes<sup>d</sup> |  |  |  |  |  |
| New or worsening SUI<sup>e</sup> | 82 | 5 (11) | 8 (22) | -11 (-30 to 5) | .23 |
| New or worsening UUI<sup>f</sup> | 82 | 8 (17) | 13 (36) | -19 (-38 to 2) | .07 |
| Treatment for UI<sup>g</sup> | 82 | 3 (7) | 7 (19) | -13 (-30 to 2) | .10 |
| SUI treatment<sup>h</sup> | 10 | 2 (67) | 1 (14) |  |  |
| UUI treatment<sup>i</sup> | 10 | 2 (67) | 7 (100) |  |  |
| Any stress urine leakage<sup>b</sup> | 82 | 2 (4) | 12 (33) | -29 (-47 to -10) | <.001 |
| Bothersome stress urine leakage<sup>b</sup> | 82 | 2 (4) | 11 (31) | -26 (-44 to -10) | .002 |
| Any urge urine leakage<sup>c</sup> | 82 | 9 (20) | 12 (33) | -14 (-34 to 6) | .20 |
| Bothersome urge urine leakage<sup>c</sup> | 82 | 8 (17) | 12 (33) | -16 (-35 to 4) | .12 |
| Urogenital Distress Inventory (UDI) score | 82 | 0.0 (0.0, 8.3) | 12.5 (0.0, 31.3) | -4.2 (-12.5 to 0.0) | .04 |
| Urinary Impact Questionnaire (UIQ) score | 82 | 0.0 (0.0, 0.0) | 0.0 (0.0, 14.3) | 0.0 (0.0 to 0.0) | .21 |
| Incontinence Severity Index (ISI) score | 82 | 0.0 (0.0, 2.0) | 1.5 (0.0, 3.5) | 0.0 (-1.0 to 0.0) | .07 |
| Change from Baseline Urinary Outcomes |  |  |  |  |  |
| Change from Baseline Urogenital Distress Inventory (UDI) score | 82 | -35.4 (-58.3, -12.5) | -16.7 (-37.5, -2.1) | -16.7 (-29.2 to -4.2) | .01 |
| Change from Baseline Urinary Impact Questionnaire (UIQ) score | 82 | -7.1 (-33.3, 0.0) | -9.5 (-31.0, 0.0) | 0.0 (-9.5 to 4.8) | .75 |
|  |  | MUS (N=46) | No MUS (N=36) |  |  |
| Change from Baseline Incontinence Severity Index (ISI) score | 82 | -3.0 (-5.0, -1.0) | 0.0 (-1.5, 1.0) | -2.0 (-3.0 to -1.0) | **<.001** |

Abbreviations: CI, confidence interval; Inf, inferior; IQR, interquartile range; ISI, Incontinence Severity Index; MRI, magnetic resonance imaging; PFDI 20, Pelvic Floor Distress Inventory 20; SUI, stress urinary incontinence; UDI, Urogenital Distress Inventory; UI, urinary incontinence; UIQ, Urinary Impact Questionnaire; UUI, urge urinary incontinence.
<sup>a</sup>For nominal categorical measures, presented as counts (percentages), the P-values were obtained from Fisher's exact test. Exact risk differences and 95% CI limits were obtained using exact methods based on the score statistic. For continuous measures, presented as medians (IQR), P-values were obtained using the Wilcoxon rank sum test. Location shift and 95% CIs were obtained using a Hodges-Lehmann estimation of location shift. All tests were conducted at the .05 significance level.
<sup>b</sup>Any stress urine leakage is defined as a positive response to PFDI-20 item 17 'Do you usually experience urine leakage related to coughing, sneezing, or laughing?' Stress urine leakage degree of bother is 0='No Leakage' for subjects with no stress urine leakage. Among subjects with any stress urine leakage, the degree of bother is reported on the follow-up question 'If yes, how much does it bother you?' corresponding to values 1='Not at all', 2='Somewhat', 3='Moderately', or 4='Quite a bit'.
<sup>c</sup>Any urge urine leakage is defined as a positive response to PFDI-20 item 16 'Do you usually experience urine leakage associated with a feeling of urgency, that is, a strong sensation of needing to go to the bathroom?' Urge urine leakage degree of bother is 0='No Leakage' for subjects with no urge urine leakage. Among subjects with any urge urine leakage, the degree of bother is reported on the follow-up question 'If yes, how much does it bother you?' corresponding to values 1='Not at all', 2='Somewhat', 3='Moderately', or 4='Quite a bit'.
<sup>d</sup>If the SUPeR visit closest to the MRI exam was at 30 or 42 months (where quality of life instruments were not collected per-protocol), patient-reported outcomes were imputed from the next closest SUPeR visit to the MRI exam with data available (i.e., 24-, 36-, or 48-month visits).
<sup>e</sup>New or worsening SUI are identified based on the collection of new or worsening SUI complication and SUI treatment<sup>g</sup> at follow-up visits 6 weeks through SUPeR visit closest to the MRI exam.
<sup>f</sup>New or worsening urge UUI are identified based on the collection of new or worsening UUI complication and UUI treatment<sup>g</sup> at follow-up visits 6 weeks through SUPeR visit closest to the MRI exam.
<sup>g</sup>Treatment for UI are identified based on the collection of UI treatment at follow-up visits 6 weeks through SUPeR visit closest to the MRI exam. Further classification of treatment for SUI and/or UUI was based on manual review by clinical expert.

Postoperatively, 13 women (16%) reported new or worsening SUI, and 21 (26%) experienced new or worsening UUI. Ten women (12%) received treatment for UI after surgery, with three (30%) having a MUS. Three women in the MUS group (7%) underwent a MUS revision. The rate of prolapse recurrence was similar between the MUS groups (MUS, 18/46 [39%] vs No-MUS, 16/36 [44%]).

Regardless of prolapse recurrence status, fewer women with MUS (vs without) experienced postoperative stress urine leakage (difference, -29%; P<.001) and bother (difference, -26%; P=.002). Across the entire cohort after surgery, the MUS group (vs No-MUS) had a lower (better) UDI score (difference, -4.2; P=.04) and showed greater improvement (larger decrease from baseline) in UDI scores (difference, -16.7; P=.01) and ISI scores (difference, -2.0; P<.001).

### Urethral versus Urinary Outcomes

Correlations between postoperative urethral measures and urinary outcomes are shown in **Supplemental Table 4**. Concerning urethral dimensions, new or worsening UUI was linked to a shorter urethral length (ρ=-0.28; P=.01) and larger proximal anterior-posterior diameter (ρ=0.31; P=.004). For urethral angles, worse postoperative UDI scores were associated with smaller urethral sagittal angles (indicating posterior deflection of the urethra away from the pubic symphysis) (ρ=-0.25; P=.02) and non-zero urethral coronal angles (representing lateral urethral inclination away from the midsagittal plane) (ρ=-0.28; P=.01). Sagittal angle correlations were related to bothersome UI leakage, while coronal angle correlations were particularly associated with UUI symptoms and bother. The statistical shape model revealed that a more “C”-shaped urethra (PC1) was linked to new or worsening UUI (ρ=-0.30; P=.006), and a more “S”-shaped urethra (PC8) correlated with poorer postoperative UIQ scores (ρ=-0.29; P=.008) (**Figure 5B/5D**).

For urethral support measures, a more posterior-inferior position of the (1) mid-to-proximal urethra correlated with worse postoperative UDI and UIQ scores (particularly new or worsening UUI), (2) mid-urethra correlated with new or worsening SUI, and (3) mid-to-distal urethra correlated with bothersome SUI leakage. Worse postoperative UIQ scores were associated with less mobility of the proximal urethra (ρ=-0.24; P=.03) and distal urethra (ρ=-0.29; P=.009).

## COMMENT

### Principal Findings

Among women without prolapse recurrence, MUS (vs no MUS) was associated with larger urethral dimensions (diameter, surface area, volume), a straighter urethral shape, better proximal urethral support (a more anterior-superior urethral position), and superior urinary outcomes, including less stress urinary leakage and bother. Poorer urinary outcomes, including postoperative SUI, were associated with a shorter urethral length, curvier urethra (more “C” or “S”-shaped urethral concavity indicated by posterior deflection of the urethral sagittal angles), and worse proximal urethral support (a more posterior-inferior urethral position).

### Results in Context of What is Known

Study findings align with previous literature showing (1) improved urethral support and urinary outcomes with MUS surgery^9^ and (2) decreased urethral support and curved urethral shape with SUI.^5^ Unlike earlier research, this study did not find urethral mobility to differ significantly with MUS^9,20^ or SUI.^12,21–24^ To our knowledge, no studies have quantified urethral shape after MUS; most have focused on urethral position and mobility rather than morphology.^9,20,25^ Discrepancies with prior research could be due to methodological differences in assessing urethral configuration (3D vs 2D).

### Clinical Implications

Our study emphasizes the significance of urethral configuration in urinary continence outcomes following prolapse repair with or without concurrent MUS. The observed relationships between postoperative urethral features, MUS, and urinary symptoms suggest that surgical placement of the MUS and urethral characteristics may affect treatment outcomes and could be optimized to improve urinary outcomes after surgery. For instance, the better urinary outcomes observed with improved urethral support from the MUS might be due to the sling acting as a backstop and encouraging scar tissue formation along the mesh after surgery, thereby further strengthening periurethral connective tissue support.^26^

Additionally, our study suggests that urethral morphology—particularly shape—may be an underappreciated factor in postoperative urinary function and urinary continence. Classical theoretical mechanisms of female urinary continence (e.g., hammock and integral theories) primarily emphasize urethral dynamics and supportive structures over the intrinsic morphology (dimensions, shape) of the urethra.^3,4^ Similarly, SUI treatment success is mainly attributed to reduced urethral mobility, optimal location of the MUS, and resolution of UI symptoms, all of which are the predominant metrics assessed in urodynamic studies of SUI and SUI repair.^11,20,26^

Consequently, urethral morphology remains largely underrepresented in urinary continence models and urodynamic studies. Similar to this work, recent studies by Routzong et al. highlight the importance of urethral morphology in female urinary continence.^5,6^ The authors proposed the “Swing Theory”, a novel theorem which explains how urethral biomechanics (dynamics, tissue properties) drive urethral shape changes (deformations) that contribute to urinary continence and the success of MUS surgery. Therefore, abnormal or suboptimal urethral morphology after surgery may contribute to postoperative UI despite anatomically successful SUI and prolapse repair.

### Research Implications

Future longitudinal studies evaluating urethral morphology alongside urethral support and urinary symptoms after prolapse and SUI repair could help towards understanding how to optimize SUI treatments. Combining imaging and clinical methods would yield outcomes that provide more insight into whether surgical approaches should be modified to better restore physiological urethral configuration and enhance urinary outcomes.

### Strengths and Limitations

Strengths of this study included its prospective design, well-defined clinical cohort, use of validated questionnaires, and 3D analysis of urethral configuration. However, there are several limitations: non-randomization of MUS placement, the limited generalizability of the homogenous cohort, the DEMAND cohort being unrepresentative of the primary SUPeR cohort, time discrepancies between MRI and urinary function data collection, no adjustments for multiple comparisons, and the absence of preoperative images to directly assess surgery-induced alterations in urethral configuration. Thus, caution is needed when interpreting study results.

### Conclusions

Urethral morphology and support vary with MUS and SUI after vaginal surgery with and without a concomitant sling. These factors were linked to postoperative urinary symptoms. Urethral configuration may impact urinary outcomes and warrant surgical consideration in prolapse and SUI repairs.

## AUTHOR CONTRIBUTIONS

M.G.G., S.T.B., and A.S. had full access to all the study data and take responsibility for its integrity and the accuracy of its analysis.

1. **S.T.B.:** Conceptualization, Data curation, Formal analysis, Visualization, Writing-review & editing, Writing-original draft.
2. **P.A.M.:** Project administration, Supervision, Conceptualization, Methodology, Funding acquisition, Formal analysis, Writing-review & editing, Writing-original draft.
3. **H.S.H.:** Conceptualization, Methodology, Funding acquisition, Writing-review & editing, Writing-original draft.
4. **C.R.R.:** Conceptualization, Methodology, Funding acquisition, Writing-review & editing, Writing-original draft.
5. **M.E.H.:** Conceptualization, Methodology, Funding acquisition, Writing-review & editing, Writing-original draft.
6. **A.C.W.:** Conceptualization, Methodology, Funding acquisition, Writing-review & editing, Writing-original draft.
7. **H.E.R.:** Conceptualization, Methodology, Funding acquisition, Writing-review & editing, Writing-original draft.
8. **T.S.G.:** Conceptualization, Methodology, Funding acquisition, Writing-review & editing, Writing-original draft.
9. **D.M.:** Project administration, Supervision, Writing-review & editing, Writing-original draft.
10. **A. S.:** Data curation, Formal analysis, Visualization, Writing-review & editing, Writing-original draft.
11. **M.G.G.:** Project administration, Supervision, Conceptualization, Methodology, Funding acquisition, Data curation, Formal analysis, Visualization, Writing-review & editing, Writing-original draft.

## DATA SHARING STATEMENT

Data from the DEMAND study will be accessible through the Eunice Kennedy Shriver National Institute of Child Health and Human Development Data and Specimen Hub (DASH) https://dash.nichd.nih.gov/ once the planned DEMAND analyses are completed (12-31-2026).

## PAPER PRESENTATION INFORMATION

This study was presented at the *American Urogynecologic Society* Pelvic Floor Disorders Week 2023, Portland, OR, USA, October 4, 2023.

## ACKNOWLEDGMENTS

In addition to the authors, the following members of the Pelvic Floor Disorders Network were involved in the Defining the Mechanisms of Anterior Vaginal Wall Descent study:

- *UC San Diego Health, San Diego, CA:* Kimberly Ferrante, Sherella Johnson, Emily S. Lukacz, Charles W. Nager.
- *Kaiser Permanente, San Diego, CA:* Gouri B. Diwadkar, Keisha Y. Dyer, Linda M. Mackinnon, Jasmine Tan-Kim, Gisselle Zazueta-Damian.
- *Duke University Medical Center, Durham, NC:* Cindy Amundsen, Yasmeen Bruton, Notorious Coleman-Taylor, Amie Kawasaki, Nicole Longoria, Shantae McLean, Nazema Siddiqui.
- *University of Alabama at Birmingham, Dept. OB/GYN, Birmingham, AL:* Kathy Carter, David Ellington, Mark E. Lockhart, Sunita Patel, Nancy Saxon, Velria B. Willis.
- *Alpert Medical School of Brown University, Providence, RI:* Cassandra Carberry, B. Star Hampton, Nicole Korbly, Ann S. Meers, Deborah L. Myers, Vivian W. Sung, Elizabeth-Ann Viscione, Kyle Wohlrab.
- *University of New Mexico:* Gena Dunivan, Yuko Komesu, Peter Jeppson.
- *University of Pennsylvania, Philadelphia, PA:* Lily Arya, Lorraine Flick, Michelle Kingslee, Ariana Smith.
- *Magee-Women’s Hospital, Dept. of OB/GYN & Reproductive Sciences, Pittsburgh, PA:* Steven D. Abramowitch, Michael Bonidie, Judy Gruss, Jonathan Shepherd, Gary Sutkin, Halina M. Zyczynski.
- *Cleveland Clinic Foundation, Cleveland, OH:* Matthew Barber, Annette Graham, Marie Fidela R. Paraiso, Cecile Ferrando.
- *Albany Medical College, Albany, NY:* Rebecca G. Rogers
- *RTI International, Research Triangle Park, NC:* Kate Burdekin, Michael Ham, Amanda Shaffer, Dennis Wallace, Ryan Whitworth, Taylor Swankie.

---

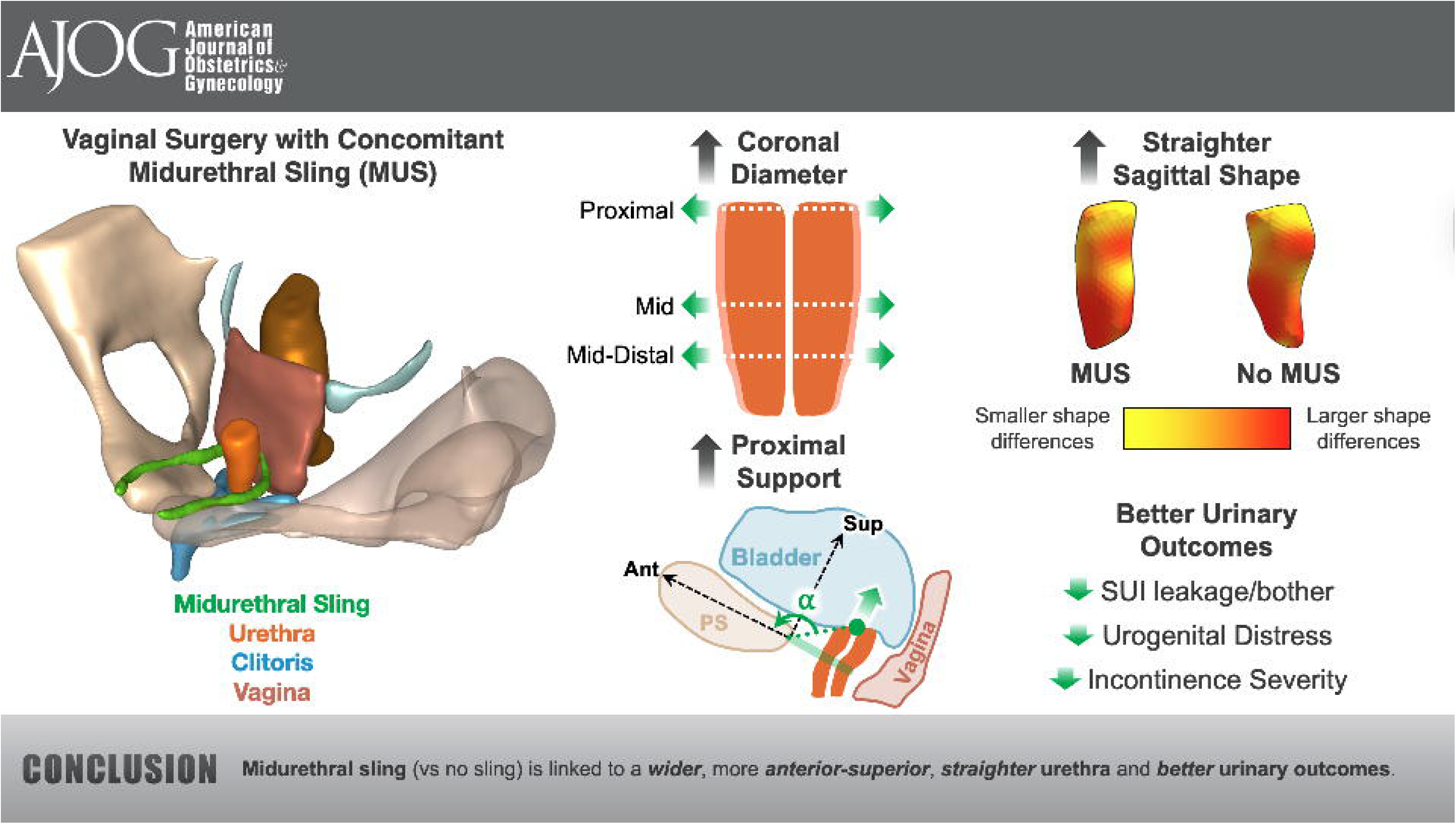

## REFERENCES

1. Giugale L, Sridhar A, Ferrante KL, et al. Long-term Urinary Outcomes After Transvaginal Uterovaginal Prolapse Repair With and Without Concomitant Midurethral Slings. Female Pelvic Med Reconstr Surg. 2022;28(3):142–148. doi:10.1097/SPV.0000000000001160

2. Wei JT, Nygaard I, Richter HE, et al. A Midurethral Sling to Reduce Incontinence after Vaginal Prolapse Repair. New England Journal of Medicine. 2012;366(25):2358–2367. doi:10.1056/nejmoa1111967

3. DeLancey JOL. Structural support of the urethra as it relates to stress urinary incontinence: the hammock hypothesis. Am J Obstet Gynecol. 1994;170(5):1713–1723.

4. Petros PEP, Ulmsten UI. An Integral Theory of Female Urinary Incontinence. Acta Obstet Gynecol Scand. 1990;69(S153):7–31. doi:10.1111/J.1600-0412.1990.TB08027.X

5. Routzong MR, Chang C, Goldberg RP, Abramowitch SD, Rostaminia G. Urethral support in female urinary continence part 1: dynamic measurements of urethral shape and motion. Int Urogynecol J. Published online 2021. doi:10.1007/s00192-021-04765-3

6. Routzong MR, Martin LC, Rostaminia G, Abramowitch S. Urethral support in female urinary continence part 2: a computational, biomechanical analysis of Valsalva. Int Urogynecol J. Published online 2021:1–11.

7. Martin LC, Routzong MR, Abramowitch SD, Rostaminia G. Effect of Squeeze, Cough, and Strain on Dynamic Urethral Function in Nulligravid Asymptomatic Women: A Cross-Sectional Cohort Study. Urogynecology. 2023;29(9):740–747. doi:10.1097/SPV.0000000000001345

8. Morgan DM, Umek W, Guire K, Morgan HK, Garabrant A, DeLancey JOL. Urethral Sphincter Morphology and Function With and Without Stress Incontinence. Journal of Urology. 2009;182(1):203-209. doi:10.1016/j.juro.2009.02.129

9. Zhao B, Zuo Y, Wen L. The relationship between urethral mobility and clinical outcomes after midurethral sling surgery. Continence. 2023;5(4):100569. doi:10.1016/j.cont.2022.100569

10. Chill H, Martin L, Abramowitch S, Rostaminia G. Effect of Mid-urethral Sling on Urethral Dynamic Shape and Motion. In: International Urogynecology Journal. Vol 33. Springer London Ltd 236 Grays Inn Rd, 6TH Floor, London WC1X 8HL, England; 2022:S292-S293.

11. Wen L, Shek KL, Subramaniam N, Friedman T, Dietz HP. Correlations between Sonographic and Urodynamic Findings after Mid Urethral Sling Surgery. Journal of Urology. 2018;199(6):1571–1576. doi:10.1016/j.juro.2017.12.046

12. Pregazzi R, Sartore A, Bortoli P, Grimaldi E, Troiano L, Guaschino S. Perineal ultrasound evaluation of urethral angle and bladder neck mobility in women with stress urinary incontinence. BJOG. 2002;109(7):821–827. doi:10.1111/j.1471-0528.2002.01163.x

13. Bowen ST, Moalli PA, Abramowitch SD, et al. Defining mechanisms of recurrence following apical prolapse repair based on imaging criteria. Am J Obstet Gynecol. Published online June 1, 2021. doi:10.1016/J.AJOG.2021.05.041

14. Nager CW, Visco AG, Richter HE, et al. Effect of vaginal mesh hysteropexy vs vaginal hysterectomy with uterosacral ligament suspension on treatment failure in women with uterovaginal prolapse: A randomized clinical trial. JAMA - Journal of the American Medical Association. 2019;322(11):1054–1065. doi:10.1001/jama.2019.12812

15. Moalli PA, Bowen ST, Abramowitch SD, et al. Methods for the Defining Mechanisms of Anterior Vaginal Wall Descent (DEMAND) Study. Int Urogynecol J. Published online September 1, 2020:1–10. doi:10.1007/s00192-020-04511-1

16. Sinex DCE, Bowen ST, Kashkoush A, et al. The establishment of a 3D anatomical coordinate system for defining vaginal axis and spatial position. Comput Methods Programs Biomed. 2021;208:106175. doi:10.1016/J.CMPB.2021.106175

17. Bowen ST, Dutta A, Rytel K, Abramowitch SD, Rogers RG, Moalli PA. 3D quantitative analysis of normal clitoral anatomy in nulliparous women by MRI. Int Urogynecol J. 2022;33(6):1649–1657. doi:10.1007/s00192-022-05172-y

18. Routzong MR, Rostaminia G, Bowen ST, Goldberg RP, Abramowitch SD. Statistical shape modeling of the pelvic floor to evaluate women with obstructed defecation symptoms. https://doi.org/101080/1025584220201813281. 2020;24(2):122-130. doi:10.1080/10255842.2020.1813281

19. Bône A, Louis M, Martin B, Durrleman S. Deformetrica 4: An Open-Source Software for Statistical Shape Analysis. In: Lecture Notes in Computer Science (Including Subseries Lecture Notes in Artificial Intelligence and Lecture Notes in Bioinformatics). 11167 LNCS. Springer Verlag; 2018:3-13. doi:10.1007/978-3-030-04747-4_1

20. Qu P, Hai N, Lv Z, Yang J. Midurethral sling position and surgical outcome: a meta-analysis. Medicine. 2024;103(2):e36115. doi:10.1097/MD.0000000000036115

21. Dietz HP, Clarke B, Herbison P. Bladder neck mobility and urethral closure pressure as predictors of genuine stress incontinence. Int Urogynecol J Pelvic Floor Dysfunct. 2002;13(5):289–293. doi:10.1007/s001920200063

22. Zhao B, Wen L, Liu D, Huang S. The Visualized Urethral Mobility Profile in Stress Urinary Incontinence Described by Four-Dimensional Transperineal Ultrasound. Journal of Ultrasound in Medicine. 2022;41(6):1439–1445. doi:10.1002/jum.15828

23. Pirpiris A, Shek KL, Dietz HP. Urethral mobility and urinary incontinence. Ultrasound in Obstetrics and Gynecology. 2010;36(4):507-511. doi:10.1002/uog.7658

24. E Jamard MBTTMRCRFAP. Utility of 2D-ultrasound in pelvic floor muscle contraction and bladder neck mobility assessment in women with urinary incontinence. J Gynecol Obstet Hum Reprod. 2019;6.

25. HS Chung SKDK. Urethral mobility and Point Aa of the Pelvic Organ Prolapse Quantification (POP-Q) system before and after midurethral sling operation. Low Urin Tract Symptoms. 2019;11(2):O117–O120.

26. Petros P. The female pelvic floor: Function, dysfunction and management according to the integral theory. The Female Pelvic Floor (Third Edition): Function, Dysfunction and Management According to the Integral Theory. Published online 2010:1–331. doi:10.1007/978-3-642-03787-0

27. Dejene SZ, Jonsson Funk M, Pate V, Wu JM. Long-term outcomes following midurethral mesh sling surgery for stress urinary incontinence. Female Pelvic Med Reconstr Surg. 2021;28(4):188. doi:10.1097/SPV.0000000000001094

