## Supplement for "Urethral Morphology and Support Associated with Urinary Symptoms after Vaginal Surgery with and without Midurethral Sling"

**SUPPLEMENTAL MATERIAL:**

**Supplemental Table 1. Inclusion and Exclusion Criteria**

| **Study Enrollment Eligibility** | **Inclusion Criteria** | **Exclusion Criteria** |
| --- | --- | --- |
| DEMAND Primary Study | - Women aged 21 or older who have completed childbearing - Prolapse beyond the hymen (defined as Ba, Bp, or C>0 cm) - Uterine descent into at least the lower half of the vagina [defined as point C>-TVL/2)] - Bothersome bulge symptoms as indicated on question 3 of the PFDI-20 form relating to ‘sensation of bulging’ or ‘something falling out’ - Desires vaginal surgical treatment for uterovaginal prolapse - Available for up to 60-month follow-up - Amenorrhea for the past 12 months from either menopause or endometrial ablation - Not pregnant, not at risk for pregnancy, or agree to contraception if at risk for pregnancy (only applicable to the rare endometrial ablation patient) - Eligible for no cervical cancer screening for at least 3 years | - Previous synthetic material (placed vaginally or abdominally) to augment POP repair - Known previous uterosacral or sacrospinous uterine suspension - Known adverse reaction to synthetic mesh or biological grafts; these complications include but are not limited to erosion, fistula, or abscess - Chronic pelvic pain - Pelvic radiation - Cervical elongation—defined as an expectation that the C point would be Stage 2 or greater postoperatively if a hysteropexy was performed (Note: cervical shortening or trachelectomy is not an allowed intraoperative procedure within the hysteropexy treatment group) - Women at increased risk of cervical dysplasia requiring cervical cancer screening more often than every 3 years [e.g., HIV+ status, immunosuppression because of transplant related medications, Diethylstilbestrol (DES) exposure in utero, or previous treatment for cervical intraepithelial neoplasia (CIN)2, CIN3, or cancer] - Uterine abnormalities (symptomatic uterine fibroids, polyps, endometrial hyperplasia, endometrial cancer, or - any uterine disease that precluded prolapse repair with uterine preservation in the opinion of the surgeon) - Indication for ovarian removal (adnexal mass, BRCA 1/2 positivity, family history of ovarian cancer) - Current condition of amenorrhea caused by exogenous sex steroids or hypothalamic conditions |
| **MRI Analysis Eligibility** | **Inclusion Criteria** | **Exclusion Criteria** |
| DEMAND Primary Study | - N/A | - Failure to capture the entire vagina - MRI taken after reoperation - Incomplete MRI |
| DEMAND Ancillary Study | - N/A | - Poor demarcation of vaginal borders |

**Supplemental Figure 1. Flow of Participants in the Defining Mechanisms of Anterior Vaginal Wall Descent (DEMAND) Ancillary Study by Concomitant Midurethral Sling**

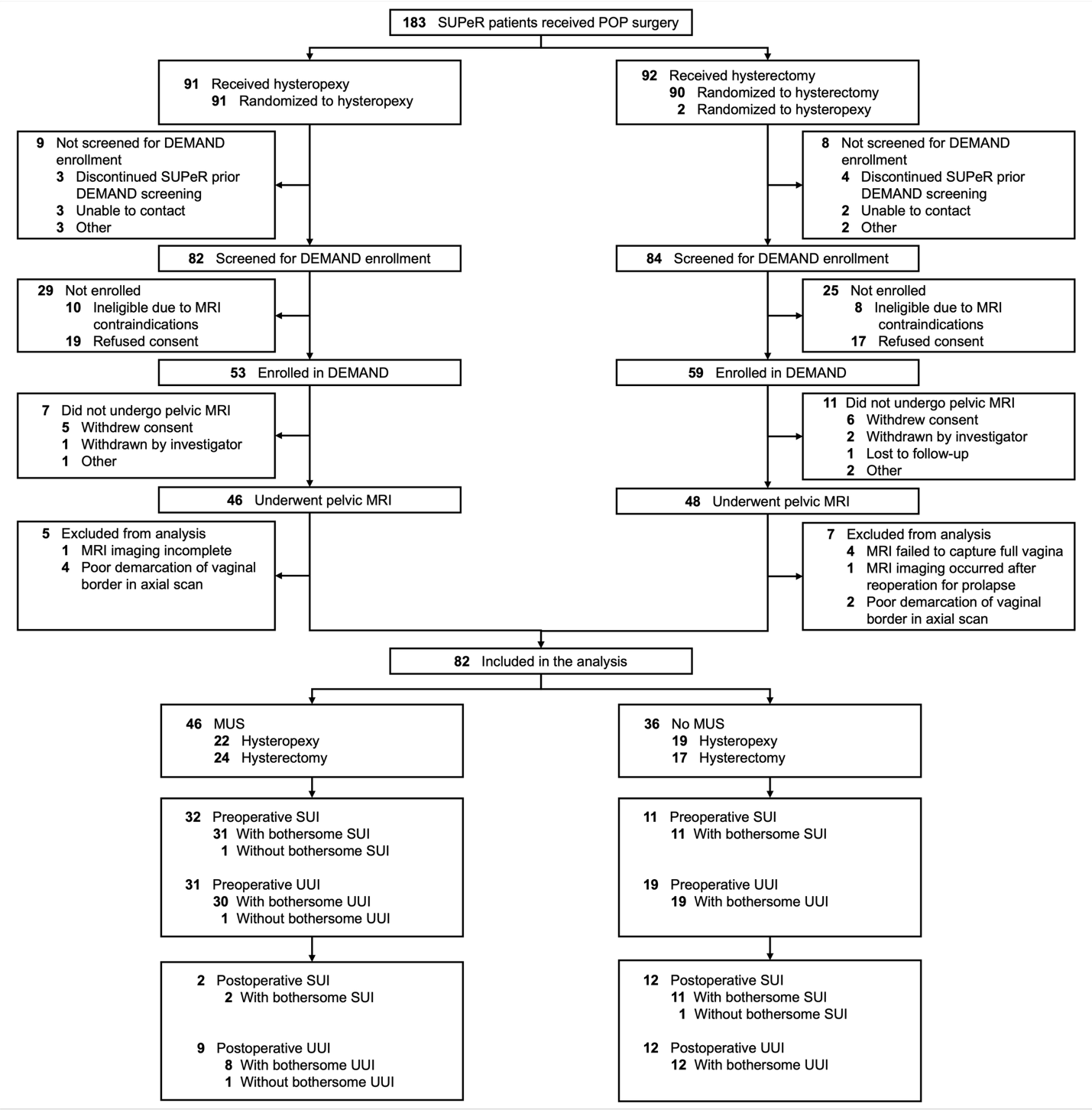

Abbreviations: DEMAND, Defining Mechanisms of Anterior Vaginal Wall Descent; MRI, magnetic resonance imaging; MUS, midurethral sling; POP, pelvic organ prolapse; SUI, stress urinary incontinence; SUPeR, Study of Uterine Prolapse Procedures-Randomized; UUI, urge urinary incontinence

Three patients—2 received hysterectomy and 1 received hysteropexy—were found ineligible for the intervention in the parent Study of Uterine Prolapse Procedures-Randomized (SUPeR) trial. Among the women who received hysterectomy, two underwent hysterectomy and sacrospinous ligament suspension.

**Supplemental Table 2. Baseline and Postoperative Demographic and Clinical Characteristics by Midurethral Sling Status**

|  |  | **Median (IQR)** | |  |  |
| --- | --- | --- | --- | --- | --- |
| **Characteristic** | **Total No.^a^** | **MUS (N=46)** | **No MUS (N=36)** | **Risk Difference/ Location Shift (95% CI)^b^** | **P-value^b^** |
| **Baseline Patient Demographics** |  |  |  |  |  |
| Age, years | 82 | 64.7 (58.6, 70.8) | 66.3 (61.7, 74.8) | -2.2 (-5.6 to 1.1) | .18 |
| White | 82 | 39 (85) | 28 (78) | 7 (-10 to 26) | .57 |
| Hispanic or Latina | 78 | 6 (14) | 3 (9) | 5 (-12 to 20) | .72 |
| Married/living with partner | 82 | 30 (65) | 22 (61) | 4 (-17 to 26) | .82 |
| Higher education after high school | 80 | 33 (73) | 22 (63) | 10 (-10 to 31) | .34 |
| Medicaid/Medicare | 82 | 21 (46) | 19 (53) | -7 (-29 to 15) | .66 |
| **Baseline Medical History** |  |  |  |  |  |
| Height (cm) | 82 | 160.0 (157.0, 165.0) | 160.0 (155.0, 168.5) | -1.0 (-5.0 to 2.0) | .47 |
| Weight (kg) | 82 | 68.0 (62.0, 74.0) | 77.5 (67.0, 84.5) | -9.0 (-13.0 to -4.0) | **.001** |
| BMI (kg/m^2^) | 82 | 25.7 (23.9, 28.9) | 29.9 (27.2, 32.3) | -3.2 (-4.9 to -1.3) | **.002** |
| Obese (BMI ≥ 30) | 82 | 10 (22) | 17 (47) | -25 (-46 to -3) | **.02** |
| Gravidity | 82 | 3.0 (2.0, 4.0) | 3.0 (2.0, 4.0) | 0.0 (0.0 to 1.0) | .64 |
| Cesarean delivery | 82 | 3 (7) | 2 (6) | 1 (-13 to 14) | >.99 |
| Vaginal parity | 82 | 3.0 (2.0, 3.0) | 2.0 (2.0, 3.0) | 0.0 (0.0 to 1.0) | .26 |
| Postmenopausal | 82 | 44 (96) | 36 (100) | -4 (-15 to 6) | .50 |
| Estrogen use | 82 | 14 (30) | 17 (47) | -17 (-38 to 5) | .17 |
| Current or historical smoker | 82 | 12 (26) | 7 (19) | 7 (-13 to 26) | .60 |
| Diabetes | 82 | 5 (11) | 4 (11) | 0 (-16 to 14) | >.99 |
| Pulmonary disease^c^ | 82 | 4 (9) | 3 (8) | 0 (-15 to 14) | >.99 |
| Cardiovascular disease^d^ | 82 | 2 (4) | 4 (11) | -7 (-22 to 6) | .40 |
| Prior POP surgery | 82 | 1 (2) | 2 (6) | -3 (-17 to 7) | .58 |
| Prior SUI surgery | 82 | 0 (0) | 5 (14) | -14 (-29 to -4) | **.01** |
| **Baseline Pelvic Floor Measurements** |  |  |  |  |  |
| POP-Q measurement (cm)^e^ |  |  |  |  |  |
| Ba | 82 | 2.0 (1.0, 4.0) | 3.0 (1.8, 4.5) | 0.0 (-1.0 to 1.0) | .54 |
| Bp | 82 | 0.3 (-1.0, 2.0) | 0.0 (-1.5, 2.0) | 0.0 (-1.0 to 1.0) | .91 |
| C | 82 | 0.0 (-3.0, 2.0) | 0.0 (-3.0, 2.0) | 0.0 (-2.0 to 1.0) | .66 |
| GH (strain) | 82 | 4.0 (4.0, 5.0) | 4.5 (3.8, 5.0) | 0.0 (-0.5 to 0.5) | .61 |
| PB (strain) | 82 | 3.0 (2.5, 3.5) | 3.0 (2.5, 4.0) | 0.0 (-0.5 to 0.0) | .85 |
| TVL | 82 | 9.0 (8.5, 10.0) | 9.0 (8.0, 10.0) | 0.0 (0.0 to 1.0) | .22 |
| Advanced POP-Q stage (≥ 3)^f^ | 82 | 32 (70) | 30 (83) | -14 (-32 to 6) | .20 |
| **Baseline Concomitant Operative Procedures** |  |  |  |  |  |
| Hysteropexy (vs hysterectomy) | 82 | 22 (48) | 19 (53) | 5 (-17 to 27) | .82 |
| Anterior prolapse repair | 82 | 36 (78) | 31 (86) | -8 (-25 to 10) | .40 |
| Posterior prolapse repair/perineorrhaphy | 82 | 27 (59) | 22 (61) | -2 (-24 to 19) | >.99 |
| **Postoperative Pelvic Floor Measurements** |  |  |  |  |  |
| POP-Q measurement (cm)^e^ |  |  |  |  |  |
| Ba | 82 | -1.0 (-2.0, 0.0) | -1.0 (-2.0, 0.0) | 0.0 (-1.0 to 0.5) | .68 |
| Bp | 82 | -1.3 (-3.0, -0.5) | -2.0 (-3.0, -1.0) | 0.5 (0.0 to 1.0) | .12 |
| C | 82 | -6.0 (-7.0, -5.0) | -6.0 (-7.0, -5.0) | 0.0 (-1.0 to 0.0) | .36 |
| GH (strain) | 82 | 3.0 (3.0, 4.0) | 3.0 (2.5, 4.0) | 0.0 (-0.5 to 0.5) | .86 |
| PB (strain) | 82 | 3.3 (3.0, 4.0) | 3.8 (3.0, 4.0) | 0.0 (-0.5 to 0.0) | .43 |
| TVL | 82 | 8.0 (7.0, 9.0) | 8.0 (7.0, 9.0) | 0.0 (-0.5 to 1.0) | .76 |
| Change from Baseline POP-Q measurement (cm)^e^ |  |  |  |  |  |
| Ba | 82 | -4.0 (-5.0, -2.0) | -4.0 (-5.3, -2.0) | 0.0 (-1.0 to 1.0) | .68 |
| Bp | 82 | -2.0 (-3.5, 0.0) | -2.0 (-4.3, 0.0) | 0.0 (-1.0 to 1.5) | .84 |
| C | 82 | -5.0 (-8.0, -3.0) | -5.5 (-7.0, -3.0) | 0.0 (-1.0 to 2.0) | .71 |
| GH (strain) | 82 | -1.0 (-1.5, -0.5) | -1.3 (-2.0, -0.3) | 0.0 (0.0 to 1.0) | .33 |
| PB (strain) | 82 | 0.3 (0.0, 1.0) | 0.5 (0.0, 1.0) | 0.0 (-0.5 to 0.0) | .64 |
| TVL | 82 | -1.0 (-2.0, 0.0) | -1.0 (-2.0, 0.0) | 0.0 (-1.0 to 0.0) | .35 |

Abbreviations: BMI, body mass index; CI, confidence interval; GH, genital hiatus; IQR, interquartile range; MRI, resonance imaging; MUS, midurethral sling; PB, perineal body; POP, pelvic organ prolapse; POP-Q, Pelvic Organ Prolapse Quantification; SUI, stress urinary incontinence; TVL, total vaginal length.

^a^A total number less than 82 indicates missing participant data.

^b^For nominal categorical measures, presented as counts (percentages), the P-values were obtained from Fisher’s exact test. Exact risk differences and 95% CI limits were obtained using exact methods based on the score statistic. For continuous measures, presented as medians (IQR), P-values were obtained using the Wilcoxon rank sum test. Location shift and 95% CIs were obtained using a Hodges-Lehmann estimation of location shift. All tests were conducted at the .05 significance level.

^c^Pulmonary disease includes any of the following: asthma, chronic obstructive pulmonary disease, acute respiratory distress syndrome, and emphysema.

^d^Cardiovascular disease includes any of the following: angina, congenital heart failure/heart disease, heart attack, stroke/transient ischemic attack, and peripheral vascular disease.

^e^POP-Q point Ba is the most distal position of the upper anterior vaginal wall. POP-Q point Bp is the most distal position of the upper posterior vaginal wall. POP-Q point C is the most distal edge of the cervix or leading edge of the vaginal cuff (hysterectomy scar). POP-Q GH is measured from the middle of the external urethral meatus to the posterior border of the hymen. POP-Q PB is measured from the posterior border of the hymen to the middle of the anal opening. POP-Q TVL is measured from the posterior fornix to the hymen when point C or D is fully reduced to its normal position.

^f^POP-Q stages: stage 2, the vagina is prolapsed between 1 cm above the hymen and 1 cm below the hymen; stage 3, the vagina is prolapsed more than 1 cm beyond the hymen but is not everted within 2 cm of its length; stage 4, the vagina is everted to within 2 cm of its length.

**Supplemental Table 3. Postoperative Urethral Measurements by Midurethral Sling Status**

|  | **Median (IQR)** | |  |  |
| --- | --- | --- | --- | --- |
| **Measurement** | **MUS (N=46)** | **No MUS (N=36)** | **Location Shift (95% CI) ^a^** | **P-value ^a^** |
| **Dimension** |  |  |  |  |
| ***Proximal Urethra*** |  |  |  |  |
| AP Diameter (mm) | 13.7 (12.7, 15.2) | 13.2 (12.3, 15.0) | 0.2 (-0.6 to 1.0) | .55 |
| Transverse Diameter (mm) | 13.9 (13.0, 14.8) | 13.3 (11.8, 15.0) | 0.5 (-0.3 to 1.4) | .22 |
| ***Mid-Proximal Urethra*** |  |  |  |  |
| AP Diameter (mm) | 14.2 (13.2, 15.9) | 14.2 (13.6, 15.3) | 0.1 (-0.7 to 0.9) | .80 |
| Transverse Diameter (mm) | 14.3 (13.5, 15.2) | 13.9 (12.9, 14.8) | 0.4 (-0.3 to 1.1) | .22 |
| ***Mid-Urethra*** |  |  |  |  |
| AP Diameter (mm) | 13.3 (12.4, 14.7) | 13.8 (12.6, 15.3) | -0.4 (-1.1 to 0.3) | .33 |
| Transverse Diameter (mm) | 13.7 (12.8, 14.6) | 13.3 (11.9, 14.3) | 0.6 (-0.2 to 1.2) | .16 |
| ***Mid-Distal Urethra*** |  |  |  |  |
| AP Diameter (mm) | 11.6 (10.7, 12.6) | 11.5 (10.8, 12.9) | 0.1 (-0.7 to 0.8) | .77 |
| Transverse Diameter (mm) | 11.9 (10.7, 13.2) | 11.4 (10.1, 12.2) | 0.7 (-0.1 to 1.5) | .09 |
| ***Distal Urethra*** |  |  |  |  |
| AP Diameter (mm) | 8.1 (7.5, 8.8) | 7.9 (7.1, 8.7) | 0.2 (-0.3 to 0.8) | .33 |
| Transverse Diameter (mm) | 7.7 (6.8, 8.5) | 7.7 (6.2, 8.5) | 0.1 (-0.5 to 0.8) | .67 |
| ***Full Urethra*** |  |  |  |  |
| Length (mm) | 32.7 (30.2, 36.1) | 33.4 (29.7, 36.5) | 0.1 (-2.0 to 2.5) | .94 |
| Surface Area (cm^2^) | 15.0 (13.5, 17.8) | 14.8 (12.8, 17.5) | 0.5 (-0.8 to 1.8) | .43 |
| Volume (cm^3^) | 4.1 (3.5, 5.4) | 4.1 (3.3, 5.1) | 0.2 (-0.3 to 0.7) | .41 |
| **Angle** |  |  |  |  |
| ***Sagittal*** |  |  |  |  |
| Proximal Angle (°)^c^ | 59.8 (48.6, 67.8) | 54.3 (46.3, 62.6) | 3.6 (-1.8 to 8.6) | .16 |
| Distal Angle (°)^c^ | 115.7 (107.0, 125.4) | 119.4 (105.0, 127.0) | -0.8 (-7.2 to 5.1) | .77 |
| Proximal-Distal Angle (°)^c^ | 175.5 (167.1, 183.4) | 171.1 (159.5, 182.3) | 3.1 (-3.9 to 10.2) | .36 |
| ***Coronal*** |  |  |  |  |
| Proximal Angle (°)^d^ | 1.4 (-1.1, 3.3) | 0.9 (-2.0, 3.7) | 0.4 (-1.5 to 2.4) | .70 |
| Distal Angle (°)^d^ | 0.7 (-1.8, 3.4) | 1.0 (-4.2, 5.0) | -0.3 (-2.6 to 2.0) | .80 |
| Proximal-Distal Angle (°)^d^ | 1.5 (-3.4, 3.9) | -0.5 (-4.5, 4.3) | 1.0 (-1.8 to 3.5) | .41 |
| **Position** |  |  |  |  |
| ***Proximal Urethra*** |  |  |  |  |
| Alpha Angle (°)^e^ | 98.6 (88.6, 108.9) | 98.2 (93.2, 117.8) | -3.6 (-12.5 to 3.4) | .36 |
| Ant-Pos Position (mm) ^f^ | -2.9 (-6.2, 0.4) | -2.9 (-8.2, -1.0) | 1.1 (-1.3 to 4.1) | .36 |
| Sup-Inf Position (mm) ^g^ | 20.9 (17.7, 24.4) | 20.7 (15.8, 23.4) | 1.1 (-1.0 to 3.4) | .29 |
| ***Mid-Proximal Urethra*** |  |  |  |  |
| Ant-Pos Position (mm) ^f^ | -6.4 (-9.2, -3.5) | -6.6 (-11.1, -4.6) | 1.1 (-0.9 to 3.3) | .29 |
| Sup-Inf Position (mm) ^g^ | 13.0 (10.5, 15.4) | 13.2 (8.4, 15.4) | 0.7 (-1.2 to 3.0) | .47 |
| ***Mid-Urethra*** |  |  |  |  |
| Ant-Pos Position (mm) ^f^ | -8.7 (-11.0, -6.8) | -8.7 (-12.1, -7.1) | 0.7 (-1.0 to 2.6) | .43 |
| Sup-Inf Position (mm) ^g^ | 5.8 (2.8, 9.4) | 5.9 (2.0, 8.0) | 1.0 (-1.2 to 3.2) | .37 |
| ***Mid-Distal Urethra*** |  |  |  |  |
| Ant-Pos Position (mm) ^f^ | -11.4 (-13.8, -10.0) | -11.2 (-15.8, -9.1) | 0.6 (-1.4 to 2.6) | .67 |
| Sup-Inf Position (mm) ^g^ | -0.5 (-4.3, 2.2) | -1.6 (-4.6, 0.9) | 1.0 (-1.1 to 3.4) | .31 |
| ***Distal Urethra*** |  |  |  |  |
| Ant-Pos Position (mm) ^f^ | -15.2 (-18.4, -12.3) | -15.6 (-20.7, -13.2) | 1.2 (-0.9 to 3.4) | .33 |
| Sup-Inf Position (mm) ^g^ | -7.5 (-12.0, -4.6) | -8.4 (-11.6, -5.9) | 0.8 (-1.4 to 3.3) | .51 |
| **Mobility** |  |  |  |  |
| Proximal Urethra Mobility (mm) | 23.9 (14.6, 31.0) | 21.2 (15.1, 29.1) | 0.3 (-4.4 to 5.7) | .91 |
| Distal Urethra Mobility (mm) | 15.4 (9.3, 18.5) | 13.8 (11.2, 17.9) | 0.5 (-3.0 to 3.0) | .73 |

Abbreviations: Ant, anterior; AP, anterior-posterior; CI, confidence interval; Inf, inferior; IQR, interquartile range; MRI, magnetic resonance imaging; MUS, midurethral sling; Pos, posterior; Sup, superior.

^a^Prolapse recurrence was defined as vaginal protrusion beyond the line between the external urethral meatus and the anterior margin of the perineal body at strain observed in the postoperative MRI.

^b^For continuous measures, presented as medians (IQR), P-values were obtained using the Wilcoxon rank sum test. Location shift and 95% CIs were obtained using a Hodges-Lehmann estimate of the location shift. All tests were conducted at the .05 significance level.

^c^Smaller values indicate a more horizontal (more posteriorly oriented) sagittal angle (towards the sacrum). Larger values indicate a more vertical (more anteriorly oriented) sagittal angle (towards the pubic symphysis).

^d^Values closer to zero indicate a more medial coronal angle (aligned with the midline). Values further away from zero indicate a more lateral coronal angle (tilted to the left or right).

^e^Larger values indicate a more posteriorly or inferiorly positioned proximal urethra/bladder neck. Smaller values indicate a more anteriorly or superiorly positioned proximal urethra/bladder neck.

^f^Larger or more positive values indicate a more anterior position. Smaller or more negative values indicate a more posterior position.

^g^Larger or more positive values indicate a more superior position. Smaller or more negative values indicate a more inferior position.

**Supplemental Table 4. Correlations between Postoperative Urethral Characteristics and Urinary Outcomes**

|  | **Correlation (P-value)^a^** | | | | | | |
| --- | --- | --- | --- | --- | --- | --- | --- |
|  | **Overall Postoperative UI** | | | **Postoperative SUI^b^** | | **Postoperative UUI^c^** | |
| **Urethral Characteristics** | **UDI Score^d^** | **UIQ Score^d^** | **ISI Score^d^** | **Leakage Degree of Bother^e^** | **New or worsening SUI^f^** | **Leakage Degree of Bother^g^** | **New or worsening UUI^h^** |
| **Dimension** |  |  |  |  |  |  |  |
| ***Proximal Urethra*** |  |  |  |  |  |  |  |
| AP Diameter | 0.17 (P=.13) | 0.06 (P=.60) | 0.10 (P=.35) | 0.05 (P=.63) | -0.06 (P=.59) | **0.31 (P=.004)** | -0.08 (P=.47) |
| Transverse Diameter | 0.14 (P=.22) | 0.09 (P=.44) | 0.13 (P=.25) | -0.07 (P=.50) | -0.13 (P=.24) | 0.18 (P=.10) | 0.00 (p>0.99) |
| ***Mid-Proximal Urethra*** |  |  |  |  |  |  |  |
| AP Diameter | 0.08 (P=.48) | -0.15 (P=.19) | 0.00 (P=.98) | 0.01 (P=.94) | 0.01 (P=.94) | 0.19 (P=.08) | -0.02 (P=.89) |
| Transverse Diameter | -0.01 (P=.92) | -0.09 (P=.41) | 0.00 (P=.98) | -0.12 (P=.29) | -0.13 (P=.24) | 0.11 (P=.31) | -0.02 (P=.83) |
| ***Mid-Urethra*** |  |  |  |  |  |  |  |
| AP Diameter | -0.05 (P=.62) | -0.17 (P=.13) | -0.08 (P=.47) | 0.04 (P=.74) | -0.04 (P=.75) | 0.07 (P=.52) | 0.00 (P=.98) |
| Transverse Diameter | -0.08 (P=.49) | -0.09 (P=.42) | -0.02 (P=.84) | -0.12 (P=.27) | -0.20 (P=.07) | 0.06 (P=.56) | -0.15 (P=.19) |
| ***Mid-Distal Urethra*** |  |  |  |  |  |  |  |
| AP Diameter | -0.10 (P=.38) | -0.06 (P=.59) | -0.03 (P=.79) | -0.12 (P=.30) | -0.12 (P=.27) | 0.04 (P=.75) | -0.02 (P=.88) |
| Transverse Diameter | -0.07 (P=.53) | -0.02 (P=.87) | 0.04 (P=.73) | -0.17 (P=.14) | -0.07 (P=.54) | 0.05 (P=.67) | -0.03 (P=.80) |
| ***Distal Urethra*** |  |  |  |  |  |  |  |
| AP Diameter | -0.06 (P=.57) | -0.02 (P=.84) | -0.04 (P=.72) | -0.17 (P=.12) | -0.04 (P=.71) | 0.06 (P=.59) | 0.19 (P=.09) |
| Transverse Diameter | -0.04 (P=.74) | 0.07 (P=.51) | 0.04 (P=.71) | -0.17 (P=.13) | -0.04 (P=.74) | 0.13 (P=.23) | 0.19 (P=.09) |
| ***Full Urethra*** |  |  |  |  |  |  |  |
| Length | -0.10 (P=.35) | -0.05 (P=.64) | 0.06 (P=.58) | 0.00 (P=.97) | -0.02 (P=.87) | -0.01 (P=.90) | **-0.28 (P=.01)** |
| Surface Area | -0.04 (P=.71) | -0.05 (P=.68) | 0.06 (P=.62) | -0.07 (P=.51) | -0.06 (P=.62) | 0.13 (P=.24) | -0.20 (P=.07) |
| Volume | -0.04 (P=.75) | -0.06 (P=.57) | 0.04 (P=.74) | -0.07 (P=.54) | -0.08 (P=.50) | 0.13 (P=.24) | -0.20 (P=.08) |
| **Angle** |  |  |  |  |  |  |  |
| ***Sagittal*** |  |  |  |  |  |  |  |
| Proximal Angle^j^ | -0.12 (P=.30) | 0.00 (p>0.99) | -0.05 (P=.68) | **-0.29 (P=.007)** | -0.11 (P=.33) | -0.05 (P=.69) | -0.01 (P=.93) |
| Distal Angle^j^ | **-0.25 (P=.02)** | **-0.23 (P=.04)** | -0.21 (P=.06) | 0.01 (P=.92) | -0.01 (P=.92) | **-0.23 (P=.03)** | -0.02 (P=.83) |
| Proximal-Distal Angle^j^ | **-0.28 (P=.01)** | -0.22 (P=.05) | -0.20 (P=.07) | -0.20 (P=.07) | -0.08 (P=.50) | **-0.25 (P=.02)** | -0.06 (P=.57) |
| ***Coronal*** |  |  |  |  |  |  |  |
| Proximal Angle^k^ | -0.17 (P=.14) | **-0.30 (P=.007)** | -0.13 (P=.25) | -0.14 (P=.22) | -0.01 (P=.91) | -0.15 (P=.18) | **-0.22 (P=.04)** |
| Distal Angle^k^ | -0.08 (P=.45) | **-0.26 (P=.02)** | -0.14 (P=.22) | -0.05 (P=.64) | -0.12 (P=.29) | **-0.25 (P=.02)** | -0.06 (P=.62) |
| Proximal-Distal Angle^k^ | -0.03 (P=.78) | -0.02 (P=.87) | 0.04 (P=.73) | -0.04 (P=.73) | 0.10 (P=.38) | 0.11 (P=.32) | -0.05 (P=.67) |
| **Position** |  |  |  |  |  |  |  |
| ***Proximal Urethra*** |  |  |  |  |  |  |  |
| Alpha Angle^l^ | 0.07 (P=.51) | -0.09 (P=.44) | -0.02 (P=.86) | **0.26 (P=.02)** | 0.13 (P=.25) | 0.00 (P=.97) | 0.10 (P=.37) |
| Ant-Pos Position^m^ | -0.04 (P=.72) | 0.13 (P=.26) | 0.06 (P=.61) | **-0.23 (P=.04)** | -0.11 (P=.33) | 0.03 (P=.77) | -0.07 (P=.52) |
| Sup-Inf Position^n^ | **-0.24 (P=.03)** | **-0.23 (P=.04)** | -0.18 (P=.11) | -0.15 (P=.19) | -0.19 (P=.09) | -0.14 (P=.22) | **-0.37 (p<.001)** |
| ***Mid-Proximal Urethra*** |  |  |  |  |  |  |  |
| Ant-Pos Position^m^ | -0.05 (P=.63) | 0.10 (P=.36) | -0.01 (P=.95) | -0.19 (P=.08) | -0.09 (P=.40) | -0.03 (P=.81) | -0.10 (P=.35) |
| Sup-Inf Position^n^ | **-0.25 (P=.03)** | **-0.24 (P=.03)** | -0.20 (P=.07) | -0.20 (P=.07) | **-0.23 (P=.04)** | -0.14 (P=.21) | **-0.34 (P=.002)** |
| ***Mid-Urethra*** |  |  |  |  |  |  |  |
| Ant-Pos Position^m^ | -0.04 (P=.73) | 0.09 (P=.41) | 0.02 (P=.89) | -0.16 (P=.15) | -0.10 (P=.37) | -0.01 (P=.93) | -0.10 (P=.36) |
| Sup-Inf Position^n^ | **-0.23 (P=.04)** | **-0.24 (P=.03)** | **-0.24 (P=.03)** | **-0.25 (P=.02)** | **-0.24 (P=.03)** | -0.15 (P=.19) | **-0.23 (P=.03)** |
| ***Mid-Distal Urethra*** |  |  |  |  |  |  |  |
| Ant-Pos Position^m^ | -0.11 (P=.31) | 0.03 (P=.82) | -0.03 (P=.78) | -0.15 (P=.17) | -0.07 (P=.56) | -0.10 (P=.36) | -0.08 (P=.46) |
| Sup-Inf Position^n^ | -0.16 (P=.16) | -0.16 (P=.16) | -0.21 (P=.06) | **-0.25 (P=.02)** | **-0.23 (P=.04)** | -0.10 (P=.39) | -0.11 (P=.33) |
| ***Distal Urethra*** |  |  |  |  |  |  |  |
| Ant-Pos Position^m^ | -0.13 (P=.25) | 0.00 (P=.98) | -0.04 (P=.70) | -0.10 (P=.35) | -0.04 (P=.69) | -0.15 (P=.18) | -0.03 (P=.82) |
| Sup-Inf Position^n^ | -0.13 (P=.26) | -0.12 (P=.27) | -0.22 (P=.05) | **-0.22 (P=.04)** | -0.18 (P=.10) | -0.07 (P=.53) | -0.06 (P=.62) |
| **Mobility** |  |  |  |  |  |  |  |
| Proximal Urethra Mobility | -0.17 (P=.13) | **-0.24 (P=.03)** | -0.15 (P=.17) | -0.01 (P=.96) | 0.02 (P=.85) | -0.02 (P=.87) | -0.09 (P=.41) |
| Distal Urethra Mobility | -0.18 (P=.11) | **-0.29 (P=.009)** | -0.17 (P=.13) | -0.06 (P=.57) | -0.18 (P=.10) | -0.09 (P=.44) | **-0.23 (P=.04)** |

Abbreviations: Ant, anterior; AP, anterior-posterior; CI, confidence interval; Inf, inferior; IQR, interquartile range; ISI, Incontinence Severity Index; MRI, magnetic resonance imaging; PFDI 20, Pelvic Floor Distress Inventory 20; Pos, posterior; SUI, stress urinary incontinence; Sup, superior; UDI, Urogenital Distress Inventory; UI, urinary incontinence; UIQ, Urinary Impact Questionnaire; UUI, urge urinary incontinence.

^a^Correlations and P-values are based on Spearman rank correlation and conducted at a 0.05 significance level.

^b^Postoperative SUI was defined as having any stress urine leakage based on PFDI-20 item 17 at the SUPeR visit closest to the MRI exam. If the SUPeR visit closest to the MRI exam was at 30 or 42 months (where quality of life instruments were not collected per-protocol), SUI status was imputed from the next closest SUPeR visit to the MRI exam with PFDI-20 item 17 data available (i.e., 24-, 36-, or 48-month visits).

^c^Postoperative UUI was defined as having any urge urine leakage based on PFDI-20 item 16 at the SUPeR visit closest to the MRI exam. If the SUPeR visit closest to the MRI exam was at 30 or 42 months (where quality of life instruments were not collected per-protocol), SUI status was imputed from the next closest SUPeR visit to the MRI exam with PFDI-20 item 17 data available (i.e., 24-, 36-, or 48-month visits).

^d^If the SUPeR visit closest to the MRI exam was at 30 or 42 months (where quality of life instruments were not collected per-protocol), patient-reported outcomes were imputed from the next closest SUPeR visit to the MRI exam with data available (i.e., 24-, 36-, or 48-month visits).

^e^Any stress urine leakage is defined as a positive response to PFDI-20 item 17 'Do you usually experience urine leakage related to coughing, sneezing, or laughing?' Stress urine leakage degree of bother is 0='No Leakage' for subjects with no stress urine leakage. Among subjects with any stress urine leakage, the degree of bother is reported on the follow-up question 'If yes, how much does it bother you?' corresponding to values 1='Not at all', 2='Somewhat', 3='Moderately', or 4='Quite a bit'.

^f^New or worsening SUI are identified based on the collection of new or worsening SUI complication and SUI treatment^i^ at follow-up visits 6 weeks through SUPeR visit closest to the MRI exam.

^g^Any urge urine leakage is defined as a positive response to PFDI-20 item 16 'Do you usually experience urine leakage associated with a feeling of urgency, that is, a strong sensation of needing to go to the bathroom?' Urge urine leakage degree of bother is 0='No Leakage' for subjects with no urge urine leakage. Among subjects with any urge urine leakage, the degree of bother is reported on the follow-up question 'If yes, how much does it bother you?' corresponding to values 1='Not at all', 2='Somewhat', 3='Moderately', or 4='Quite a bit'.

^h^New or worsening UUI are identified based on the collection of new or worsening UUI complication and UUI treatment^i^ at follow-up visits 6 weeks through SUPeR visit closest to the MRI exam.

^i^Treatment for UI are identified based on the collection of UI treatment at follow-up visits 6 weeks through SUPeR visit closest to the MRI exam. Further classification of treatment for SUI and/or UUI was based on manual review by clinical expert.

^j^Smaller values indicate a more horizontal (more posteriorly oriented) sagittal angle (towards the sacrum). Larger values indicate a more vertical (more anteriorly oriented) sagittal angle (towards the pubic symphysis).

^k^Values closer to zero indicate a more medial coronal angle (aligned with the midline). Values further away from zero indicate a more lateral coronal angle (tilted to the left or right).

^l^Larger values indicate a more posteriorly or inferiorly positioned proximal urethra/bladder neck. Smaller values indicate a more anteriorly or superiorly positioned proximal urethra/bladder neck.

^m^Larger or more positive values indicate a more anterior position. Smaller or more negative values indicate a more posterior position.

^n^Larger or more positive values indicate a more superior position. Smaller or more negative values indicate a more inferior position.
